# TNFα-producing CD4^+^ T cells dominate the SARS-CoV-2-specific T cell response in COVID-19 outpatients and are associated with durable antibodies

**DOI:** 10.1101/2022.01.25.22269670

**Authors:** Kattria van der Ploeg, Adam S. Kirosingh, Diego A. M. Mori, Saborni Chakraborty, Zicheng Hu, Benjamin L. Seivers, Karen B. Jacobson, Hector Bonilla, Julie Parsonnet, Jason R. Andrews, Kathleen D. Press, Maureen C. Ty, Daniel R. Ruiz-Betancourt, Lauren de la Parte, Gene S. Tan, Catherine A. Blish, Saki Takahashi, Isabel Rodriguez-Barraquer, Bryan Greenhouse, Upinder Singh, Taia T. Wang, Prasanna Jagannathan

## Abstract

SARS-CoV-2-specific CD4^+^ T cells are likely important in immunity against COVID-19, but our understanding of CD4^+^ longitudinal dynamics following infection and specific features that correlate with the maintenance of neutralizing antibodies remains limited. We characterized SARS-CoV-2-specific CD4^+^ T cells in a longitudinal cohort of 109 COVID-19 outpatients. The quality of the SARS-CoV-2-specific CD4^+^ response shifted from cells producing IFNγ to TNFα^+^ from five days to four months post-enrollment, with IFNγ^-^IL21^-^TNFα^+^ CD4^+^ T cells the predominant population detected at later timepoints. Greater percentages of IFNγ^-^IL21^-^TNFα^+^ CD4^+^ T cells on day 28 correlated with SARS-CoV-2 neutralizing antibodies measured seven months post-infection (ρ=0.4, P=0.01). mRNA vaccination following SARS-CoV-2 infection boosted both IFNγ and TNFα producing, spike protein-specific CD4^+^ T cells. These data suggest that SARS-CoV-2-specific, TNFα-producing CD4^+^ T cells may play an important role in antibody maintenance following COVID-19.

## Introduction

Severe acute respiratory syndrome coronavirus 2 (SARS-CoV-2) has caused hundreds of millions of cases of coronavirus disease 2019 (COVID-19) worldwide, resulting in more than 5 million deaths (*Mortality Analyses*, 2021). Following primary infection or vaccination, individuals generate an adaptive immune response consisting of SARS-CoV-2-specific antibodies, B cells, and T cells; the emergence of this response is associated with successful resolution of COVID-19 symptoms (Chen & John Wherry, 2020; Grifoni et al., 2020; Sette & Crotty, 2021). This adaptive immune memory response is likely also critical in protecting individuals against re-infection. Following both infection and vaccination, waning titers of neutralizing antibodies strongly correlate with a risk of re-infection (Anand et al., 2021; Cromer et al., 2021). Although several studies have identified long-lived memory immune responses in individuals following infection and vaccination (Breton et al., 2021; Dan et al., 2021; Goel et al., 2021; Painter et al., 2021; Rodda et al., 2021; Tan et al., 2021; Wheatley et al., 2021; Zuo et al., 2021), more comprehensive, longitudinal analyses are limited (Cohen et al., 2021; Jung et al., 2021). A better understanding of how the quality of SARS-CoV-2-specific memory immune responses change over time, and which features correlate with durable antibodies, are key towards understanding long-lived immunity to SARS-CoV-2.

Most COVID-19 vaccine development has focused on the generation of neutralizing SARS-CoV-2-specific antibodies (Corey et al., 2020; Thanh Le et al., 2020), given their clinical utility when given via passive transfer (Weinreich et al., 2021) and association with protection against infection (Addetia et al., 2020; Huang et al., 2020; Khoury et al., 2021). SARS-CoV-2-specific CD4^+^ T cell responses are likely critical to the generation of these antibodies, since long-term humoral immunity is dependent on CD4^+^ T cell help (Crotty, 2019; Hale & Ahmed, 2015). SARS-CoV-2-specific CD4^+^ T cell responses are more dominant than CD8^+^ T cell responses (Grifoni et al., 2020; Sekine et al., 2020; Zuo et al., 2021), and have been associated with milder disease in acute and convalescent COVID-19 patients (Liao et al., 2020; Rydyznski Moderbacher et al., 2020; Sekine et al., 2020; Zhou et al., 2020), suggesting that this response may also play an important role in controlling and resolving a primary SARS-CoV-2 infection. SARS-CoV-2-specific CD4^+^ Th1 cells, producing interferon-gamma (IFNγ), tumor necrosis factor alpha (TNFα), and/or interleukin (IL)2, have been identified (Braun et al., 2020; Grifoni et al., 2020; Peng et al., 2020; Rydyznski Moderbacher et al., 2020; Sekine et al., 2020; Weiskopf et al., 2020), suggesting an important role for polyfunctional Th1 cells in the antiviral response, analogous to other viral infectious diseases (Seder et al., 2008). Besides Th1 cells, virus-specific CD4^+^ T cells also differentiate into T follicular helper (Tfh) cells to instruct B cells, by producing IL21 for instance, to develop long-term humoral immunity (Crotty, 2019; Eto et al., 2011; Hale & Ahmed, 2015; Rasheed et al., 2013). Circulating Tfh cells (cTfh) have been identified following acute SARS-CoV-2 infection (Boppana et al., 2021; Gong et al., 2020; Juno et al., 2020; Meckiff et al., 2020; Neidleman et al., 2020; Rodda et al., 2021; Rydyznski Moderbacher et al., 2020; Sekine et al., 2020; Wheatley et al., 2021), although their formation may be delayed (Boppana et al., 2021), and their relationship with the antibody response remains unclear. Studying the relationship between different CD4^+^ T cell responses detected following infection, and which responses most strongly correlate with maintenance of antibody responses following infection, is critical towards the development of next generation vaccines.

To date, most SARS-CoV-2-specific T cells studies have been limited by either small sample size, assessing acute and convalescent samples in different individuals, and/or evaluating correlations between SARS-CoV-2-specific CD4^+^ T cell responses and SARS-CoV-2-specific antibody titers measured at concurrent, rather than prospective, timepoints. To better evaluate the quality of the antigen-specific CD4^+^ T cell response following infection, and determine whether antigen-specific cTfh or other Th populations correlate with durable neutralizing antibodies following infection, we characterized the SARS-CoV-2-specific T cell response over time in a longitudinal cohort of 109 COVID-19 outpatients. Individuals were enrolled within 3 days of PCR-based diagnosis and sampled repeatedly at multiple timepoints out to ten months post-enrollment. Moreover, a subset of participants received both doses of mRNA vaccination during follow-up, enabling us to evaluate SARS-CoV-2-specific T cell responses in previously infected individuals following vaccination. Our data provide insight into the shifting quality of the SARS-CoV-2-specific CD4^+^ T cell response following infection and how this is impacted by vaccination. We further identify SARS-CoV-2-specific CD4^+^ T cell features that are most strongly correlated with neutralizing antibodies. Collectively, our data suggest an important role of SARS-CoV-2-specific, TNFα-producing CD4^+^ T cells in antibody maintenance following COVID-19.

## Results

### SARS-CoV-2-specific CD4^+^ T cells produce IFNγ, TNFα, and IL21 alone or in combination, with TNFα producing cells the dominant population

PBMCs were obtained from 109 COVID-19 outpatients enrolled in a Phase 2 trial of Peginterferon Lambda-1a at enrollment (D0), and 5, 14, 28, and 120 days post-enrollment. The median age of participants was 37 years, 59% percent were male, and the median duration of symptoms prior to enrollment was five days (IQR 4-7 days, Supplemental Table 1). Following enrollment, participants completed a daily at home symptom questionnaire, with a median duration until symptom resolution of eight days following enrollment (Jagannathan et al., 2021). As previously described, two participant groups were identified based on symptom trajectory analysis, with one cluster (n=7/109 (6.4%)) characterized by greater symptom severity and/or later peak severity, especially chest pain/pressure, fatigue, and myalgias, and the other cluster characterized by more mild symptoms (Jacobson et al., 2022).

We first investigated the magnitude and quality of SARS-CoV-2-specific T cell responses on day 28 (D28) post-enrollment using intracellular cytokine staining (ICS). PBMCs were stimulated with two PepTivator SARS-CoV-2 peptide pools: membrane glycoprotein and nucleocapsid phosphoprotein, pooled together (MN), or spike (S) protein, and assessed for intracellular production of IFNγ, TNFα, IL10, and IL21. Antigen-specific CD4^+^ T cells were identified by first gating on non-naïve CD4^+^ T cells (excluding CCR7^+^CD45RA^+^ CD4^+^ T cells, Figure 1A and Supplemental Figure 1A). As we detected no antigen-specific CD4^+^ T cell production of IL10 (Supplemental Figure 1C), downstream analyses focused on IFNγ, TNFα, and IL21. We found that SARS-CoV-2-specific CD4^+^ T cells consist of four distinct cytokine-producing populations: TNFα alone (IFNγ^-^IL21^-^TNFα^+^), IFNγ alone (IFNγ^+^IL21^-^TNFα^-^), IL21 alone (IFNγ^-^IL21^+^TNFα^-^) or both IFNγ and TNFα (IFNγ^+^IL21^-^TNFα^+^), with the highest percentages of cells being IFNγ^-^IL21^-^TNFα^+^ (Figure 1B and Supplemental Figure 1C). In separate experiments, we found that the majority of IFNγ^-^IL21^-^TNFα^+^-and IFNγ^+^IL21^-^TNFα^+^ CD4^+^ T cells also co-produce IL2 (Supplemental Figure 2). Percentages of MN-specific IFNγ^-^ IL21^-^TNFα^+^, IFNγ^+^IL21^-^TNFα^-^ and IFNγ^+^IL21^-^TNFα^+^ cells were slightly greater compared to S protein-specific cells (Figure 1B). When considering the quality of the SARS-CoV-2 specific CD4^+^ T cell response (e.g., the proportion of antigen-specific cells making specific cytokines, alone or in combination), >50% of the MN-and S-specific CD4^+^ T cell response was comprised of IFNγ^-^IL21^-^TNFα^+^ cells (Figure 1B). CD4^+^ T cells producing TNFα alone were significantly higher in males compared to females, older participants (>35 years of age), and those experiencing greater symptom severity and/or later peak severity as previously defined (Jacobson et al., 2022) (Supplemental Figure 3), consistent with published reports (Cohen et al., 2021; Dan et al., 2021). No significant differences in cytokine-producing SARS-CoV-2-specific CD4^+^ T cells were observed between Peginterferon Lambda and placebo treatment arms (Supplemental Figure 4A,4 B). Together, these data suggest that SARS-CoV-specific IFNγ^-^ IL21^-^TNFα^+^ cells are the dominant population detected 28 days post-enrollment in this outpatient cohort, and their magnitude is influenced by participant gender, age, and disease severity.

**Figure 1.**
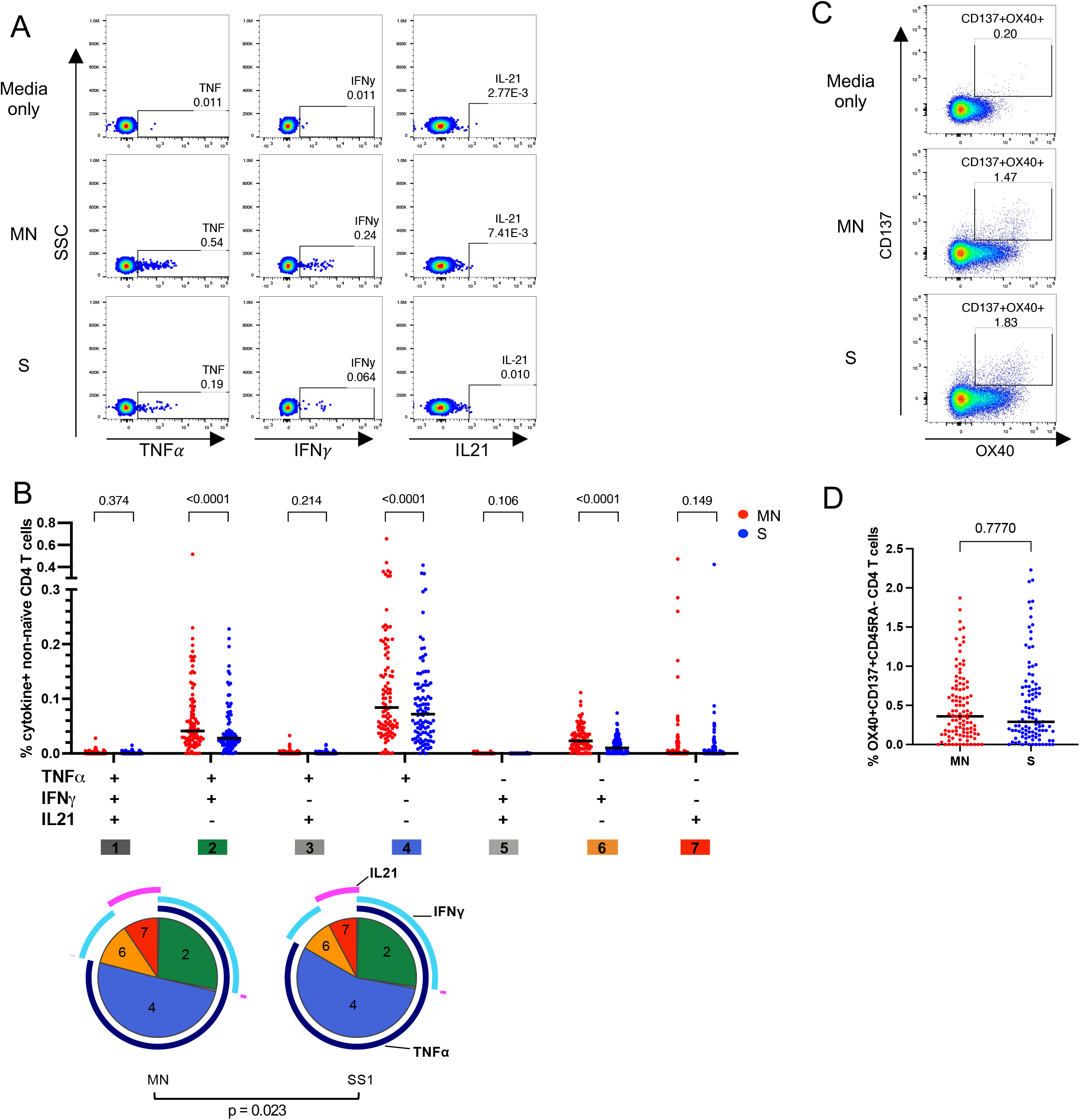
Identification of four distinct SARS-CoV-2-specific CD4^+^ T cell subsets that differ between S and MN protein stimulation. PBMCs of COVID-19 outpatients on D28 post-enrollment were stimulated with spike (S) or combination of membrane (M) and nucleocapsid (N) proteins in vitro, stained and analyzed by flow cytometry. Representative flow plots of A) cytokine-producing (TNFα, IFNγ and IL21) or C) AIM-expressing (CD137^+^OX40^+^) CD45RA^-^ CD4^+^ T cells that are stimulated with ‘media only’, MN or S proteins. B) The absolute percentage (scatter plot, black line = median) of each individual combination of TNFα, IFNγ and IL21^-^producing non-naïve CD4^+^ T cells stratified by antigenic stimuli (red: MN, blue: S, n=99). The pie charts represent the relative proportion (mean) of each individual combination of cytokine-producing non-naïve CD4^+^ T cells are shown stratified by antigenic stimuli (MN: n=102, S: n=100). The P value shown under the pie charts is calculated using a partial permutation test. D) Shown are absolute percentages of OX40^+^CD137^+^ CD45RA^-^ CD4^+^ T cells stratified by antigenic stimuli with the black line indicating the median (n=104). B, D) P values shown above the scatter plots are calculated using Wilcoxon matched-pairs signed rank test. Values shown are background (’media only’ condition) subtracted.

As an alternate approach to ICS, we also performed an activation induced marker (AIM) assay to assess SARS-CoV-2-specific CD4^+^ T cells via upregulation of CD137 and OX40 (Supplemental Figure 1C) (Dan et al., 2021; Grifoni et al., 2020; Reiss et al., 2017; Rydyznski Moderbacher et al., 2020). In contrast to responses characterized by ICS, we observed similar percentages of AIM^+^ CD4^+^ T cells following MN-and S-protein stimulation (Figure 1D). In addition, SARS-CoV-2-specific CD4^+^ T cells were more abundant compared to SARS-CoV-2-specific CD8^+^ T cells (Supplemental Figure 1C), consistent with other reports (Grifoni et al., 2020; Sekine et al., 2020; Zuo et al., 2021). As with the ICS assay, responses were similar between Lambda and placebo treatment arms in SARS-CoV-2-specific AIM^+^ CD4^+^ and CD8^+^ T cell responses, and in SARS-CoV-2 full length S binding IgG or neutralizing antibody titers measured on D28 post-enrollment (Supplemental Figure 4C-E). These data suggest that a single dose of Peginterferon Lambda treatment did not have a measurable impact on the adaptive immune response 28 days post-treatment.

### Longitudinal analysis suggests a shift in quality of SARS-CoV-2-specific T cell response from predominantly IFNγ to TNFα over time

We next investigated the magnitude and quality of the CD4^+^ T cell response by ICS over time, starting from the day of enrollment (D0) until month four (D120) post-enrollment in the same individuals. On day five (D5) SARS-CoV-2-specific CD4^+^ T cells primarily produced IFNγ (Figure 2A, 2B and Supplemental Figure 5A). At day 14 (D14), we observed a shift in the quality of the MN and S protein-specific T cell response, with significant increases in the magnitude and proportion of IFNγ^-^IL21^-^TNFα^+^ and IFNγ^+^IL21^-^TNFα^+^ CD4^+^ T cells. These TNFα-producing CD4^+^ T cells were the predominant population of SARS-CoV-2-specific CD4^+^ detected up to four months after enrollment, while the percentage of IFNγ^-^producing CD4^+^ T cells declined over time. The magnitude of SARS-CoV-2-specific IFNγ^-^IL21^+^TNFα^-^ CD4^+^ T cells was low (Figure 2A, 2B and Supplemental Figure 5A). This changing quality of CD4^+^ T cells was observed following both MN and S protein stimulation (Figure 2B). Similar results were observed when analyzing T cell responses considering days since symptom onset (rather than days since enrollment) (Supplemental Figure 5B-D), as well as when analyzing the samples from the placebo arm only (Supplemental Figure 4F).

**Figure 2.**
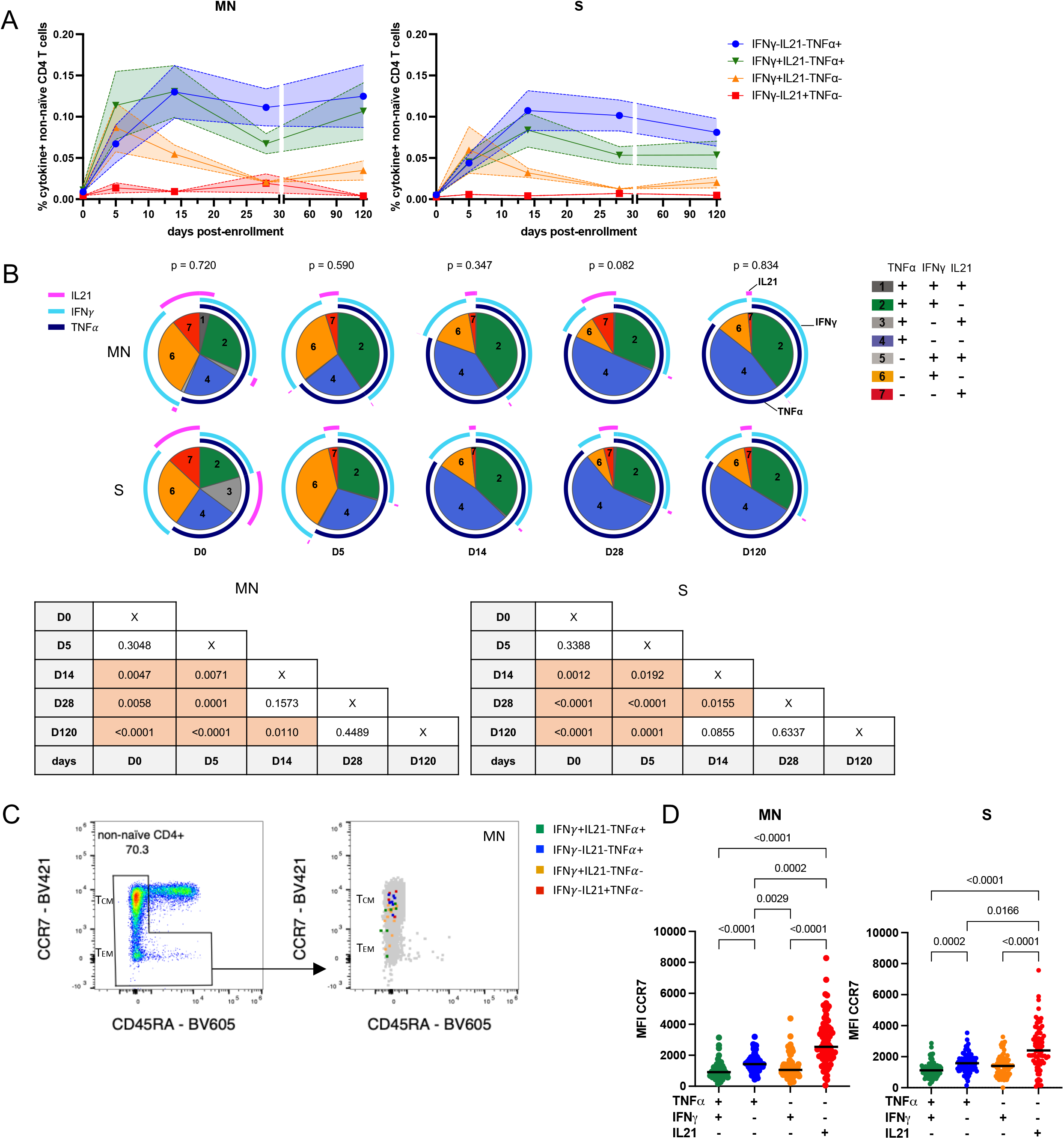
Kinetics of four distinct cytokine-producing SARS-CoV-2-specific CD4^+^ T cell populations over time and the density of CCR7 receptor expression on these populations. A,B) PBMCs from 24 COVID-19 outpatients sampled at day 5 (D5), day 14 (D14), day 28 (D28) and month 4 (D120) (n=10 also sampled on day of enrollment (D0)) were stimulated with MN (left) or S (right) proteins in vitro, stained and analyzed by flow cytometry. A) The mean and SEM of the absolute percentage of background-subtracted IFNγ^-^IL21^-^TNFα^+^- (blue), IFNγ^+^IL21^-^TNFα^-^- (yellow), IFNγ^-^IL21^+^TNFα^-^- (red) or IFNγ^+^IL21^-^TNFα^+^-producing non-naïve CD4^+^ T cells (green) of sequential timepoints post-enrollment are shown. These four populations were the dominant populations identified in Figure 1. B) The relative proportion of each individual combination of background-subtracted, TNFα, IFNγ and IL21^-^producing non-naïve CD4^+^ T cells stratified by sequential days are depicted in the pie charts (mean). The P values indicated on top of the pie charts are calculated using the partial permutation test which tests the association between MN and S protein stimulation of each indicated day post-enrollment. The P values in the tables indicate the significance of the associations between the different days post-enrollment calculated using the partial permutation test (left = MN protein-stimulated T cells, right = S protein-stimulated T cells). C) Representative flow cytometry plots of CCR7 and CD45RA-expressing CD4^+^ T cells (left) with an overlay of MN protein-specific IFNγ^-^IL21^-^ TNFα^+^- (blue), IFNγ^+^IL21^-^TNFα^-^- (yellow), IFNγ^-^IL21^+^TNFα^−^-(red) or IFNγ^+^IL21^-^TNFα^+^- producing non-naïve CD4^+^ T cells (green, right). D) MFI of CCR7 on MN (left, n=72) or S protein-specific (right, n=74) single positive TNFα- (blue), IFNγ^-^ (yellow), IL21^-^ (red) or TNFα and IFNγ^-^producing non-naïve CD4^+^ T cells (green) from day 28 post-enrollment. The P values were calculated using the Friedman test with Dunn’s multiple comparisons test (black line = median).

To further characterize these IFNγ^+^ and TNFα^+^ cells, we examined CCR7 cell surface receptor expression to better understand their differentiation status (e.g., central memory versus effector memory). SARS-CoV-2-specific IFNγ^-^IL21^-^TNFα^+^ CD4^+^ T cells had significantly higher CCR7 mean fluorescence intensity compared to IFNγ^+^IL21^-^TNFα^-^ or IFNγ^+^IL21^-^TNFα^+^ cells (Figure 2C, 2D). This suggests that CD4^+^ T cells producing TNFα alone were more central memory-like, while the IFNγ^-^producing populations were more effector memory-like. Together, these data suggest a shift in the magnitude and quality of the SARS-CoV-2-specific CD4^+^ T cell response over time with a switch from an IFNγ^-^producing effector memory-like response at early timepoints to a TNFα-producing, central memory-like response at later timepoints.

### Activated cTfh, SARS-CoV-2-specific cTfh cell and AIM^+^ CD4^+^ T cell percentages correlate with SARS-CoV-2-specific TNFα and IFNγ cytokine-producing CD4^+^ T cells

We next profiled other CD4^+^ T cell populations longitudinally, including SARS-CoV-2-specific AIM^+^ (CD137^+^OX40^+^) CD4^+^, cTfh (PD-1^+^CXCR5^+^CD4^+^), activated cTfh (ICOS^+^), and SARS-CoV-2-specific AIM^+^ cTfh cells (PD-1^+^CXCR5^+^OX40^+^CD137^+^) (Figure 3B and Supplemental Figure 1B for gating). We observed similar percentages of SARS-CoV-2-specific AIM^+^ CD4^+^ T cells between D5 and D28, although both MN and S protein-specific AIM^+^ CD4^+^ declined between D28 and D120 (Figure 3A). In general, cTfh percentage remained similar over time with the exception of a greater percentage of cTfh cells on D5 compared to D28 (Figure 3C). Activated cTfh percentage declined after D14 (Figure 3D). SARS-CoV-2-specific AIM^+^ Tfh cells were detectable up to four months after enrollment and magnitudes remained largely unchanged over time (Figure 3E).

**Figure 3.**
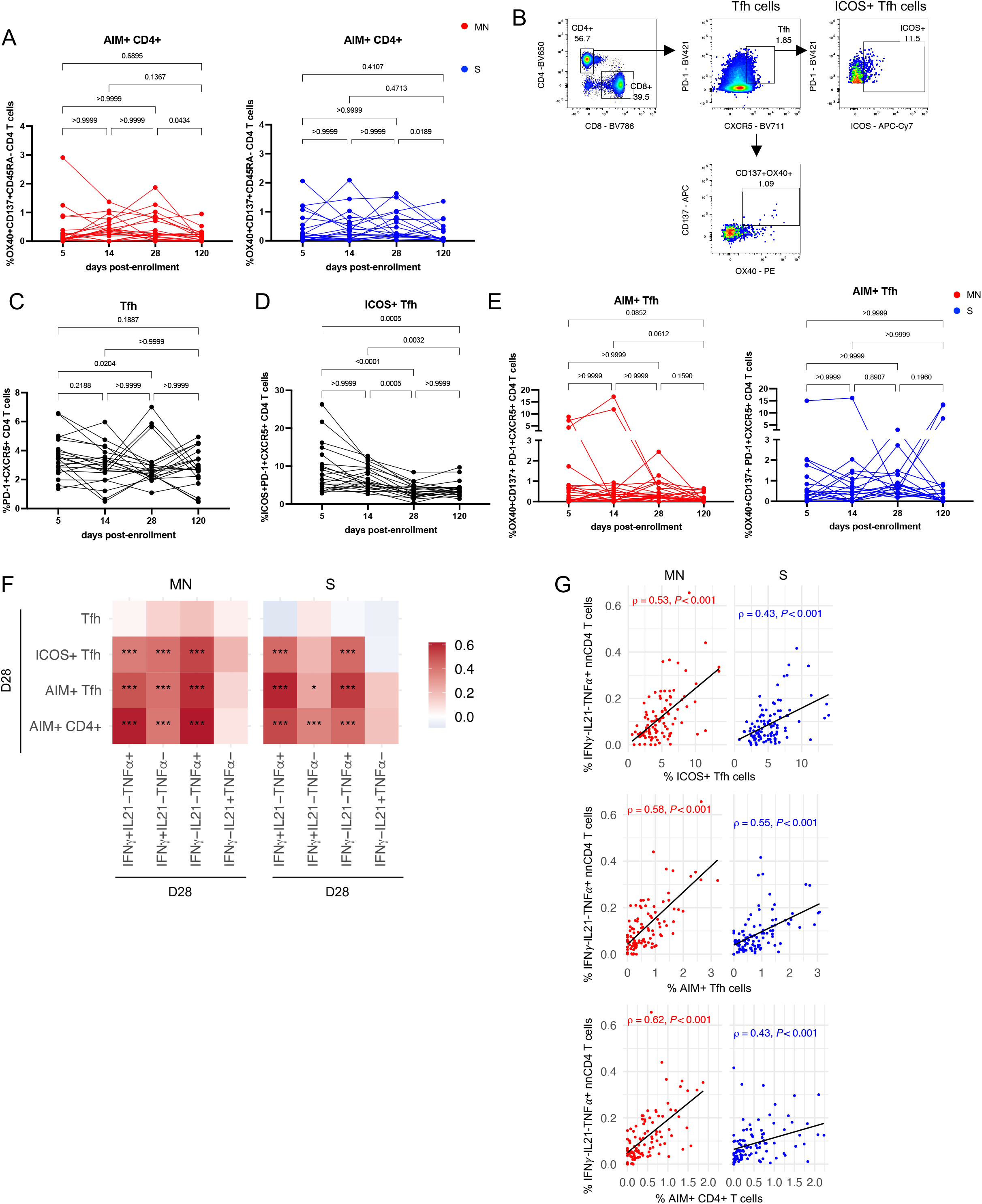
The kinetics of SARS-CoV-2-specific AIM^+^ CD4^+^, activated cTfh and SARS-CoV-2-specific cTfh cell percentages and their correlation with cytokine-producing CD4^+^ T cells. PBMCs of COVID-19 outpatients on day 5, 14, 28 and 120 post-enrollment were stimulated with MN or S proteins in vitro, stained and analyzed by flow cytometry. A) The kinetics of the absolute percentage of paired MN (left, n=22) or S (right, n=19) protein-stimulated AIM^+^ CD45RA^-^ CD4^+^ T cells over time. B) The gating strategy of PD-1^+^CXCR5^+^CD4^+^ (cTfh), PD-1^+^CXCR5^+^ICOS^+^CD4^+^ (ICOS^+^ cTfh) and PD-1^+^CXCR5^+^OX40^+^CD137^+^CD4^+^ (AIM^+^ cTfh) cells. The kinetics of the absolute percentage of paired C) cTfh (n=21) and D) ICOS^+^ cTfh (n=21) cells of unstimulated cells (‘media only’) are depicted. E) The kinetics of the absolute percentage paired AIM^+^ cTfh cells stimulated with MN (left, n=22) or S (right, n=19) protein over time are shown. A,C-E) The P values were calculated using the Friedman test with Dunn’s multiple comparisons test. F) The heatmap shown depicts spearman’s correlations (Benjamini-Hochberg corrected) between the percentages of MN protein-(left, n=101) or S protein-stimulated (right, n=100) cTfh cell populations (cTfh, ICOS^+^ cTfh and AIM^+^ cTfh) or AIM^+^ CD45RA^-^ CD4^+^ T cells measured in AIM experiments and cytokine-producing non-naïve CD4^+^ T cells measured in ICS experiments. The measurements that are significantly associated are indicated by asterisks (*=P<0.05, ***=P<0.001). G) Scatter plots comparing antigen-stimulated (MN: left/red, n=101, S: right/blue, n=100) IFNγ^-^IL21^-^ TNFα^+^-producing T cells with ICOS^+^ cTfh (top), AIM^+^ cTfh (middle) or AIM^+^ CD45RA^-^ CD4^+^ T (bottom) cells are shown (n=101). The rho (ρ) and P values were calculated using spearman’s (Benjamini-Hochberg corrected). The lines represent the fitted linear relationship between the indicated cell populations.

Subsequently, we assessed the relationship between production of cytokines and expression of activation markers, specifically investigating correlations between 1) SARS-CoV-2-specific cytokine-producing CD4^+^ T cell percentages measured in ICS experiments and 2) Percentages of SARS-CoV-2-specific AIM^+^ CD4^+^ and the three different cTfh populations measured in the AIM assays, since it is unclear how these T cell populations relate to each other. All IFNγ^-^and TNFα-producing CD4^+^ T cell populations positively and significantly correlated with AIM^+^ CD4^+^, AIM^+^ cTfh, and ICOS^+^ cTfh. In contrast, IFNγ^-^IL21^+^TNFα^-^ CD4^+^ T cells did not correlate with circulating percentages of cTfh cells (Figure 3F, 3G). Together, these data suggest that SARS-CoV-2-specific TNFα and IFNγ^-^producing CD4^+^ T cell populations highly correlated with SARS-CoV-2-specific AIM^+^ CD4^+^ T cell and cTfh cell populations, but that, in our assay, IL21 production did not correlate with either AIM^+^ or cTfh cell populations.

### SARS-CoV-2-specific TNFα^+^ CD4 T cell and activated Tfh cell responses on D28 correlate with durable antibody responses

Neutralizing antibodies are thought to be the primary adaptive effector function that mediate protection from COVID-19 re-infection (Addetia et al., 2020; Huang et al., 2020; Khoury et al., 2021). CD4^+^ T helper responses are important in germinal center immune reactions and affinity maturation, and antigen-specific cTfh were recently described to be a correlate of the peak antibody response following SARS-CoV-2 vaccination (Painter et al., 2021). We thus explored whether specific CD4^+^ T cell features following natural infection correlated with the antibody response measured at timepoints in the future, including S protein binding IgG and neutralizing antibody titers (Chakraborty et al., 2021).

We first evaluated whether SARS-CoV-2-specific CD4^+^ T cell responses measured early following infection (D5) correlated with the peak antibody response, measured on D28 post-enrollment (Chakraborty et al., 2021; Wei et al., 2021). MN protein-specific, IFNγ^+^-producing responses measured on D5 positively correlated with S protein binding IgG on D28, although this correlation was not significant after multiple comparison correction (Figure 4A, 4B, Supplemental Figure 6). Furthermore, percentages of antigen-specific cTfh, and other CD4^+^ T cell populations, measured on D5 were also not significantly correlated with SARS-CoV-2-specific antibody responses measured on D28 (Figure 4A).

**Figure 4.**
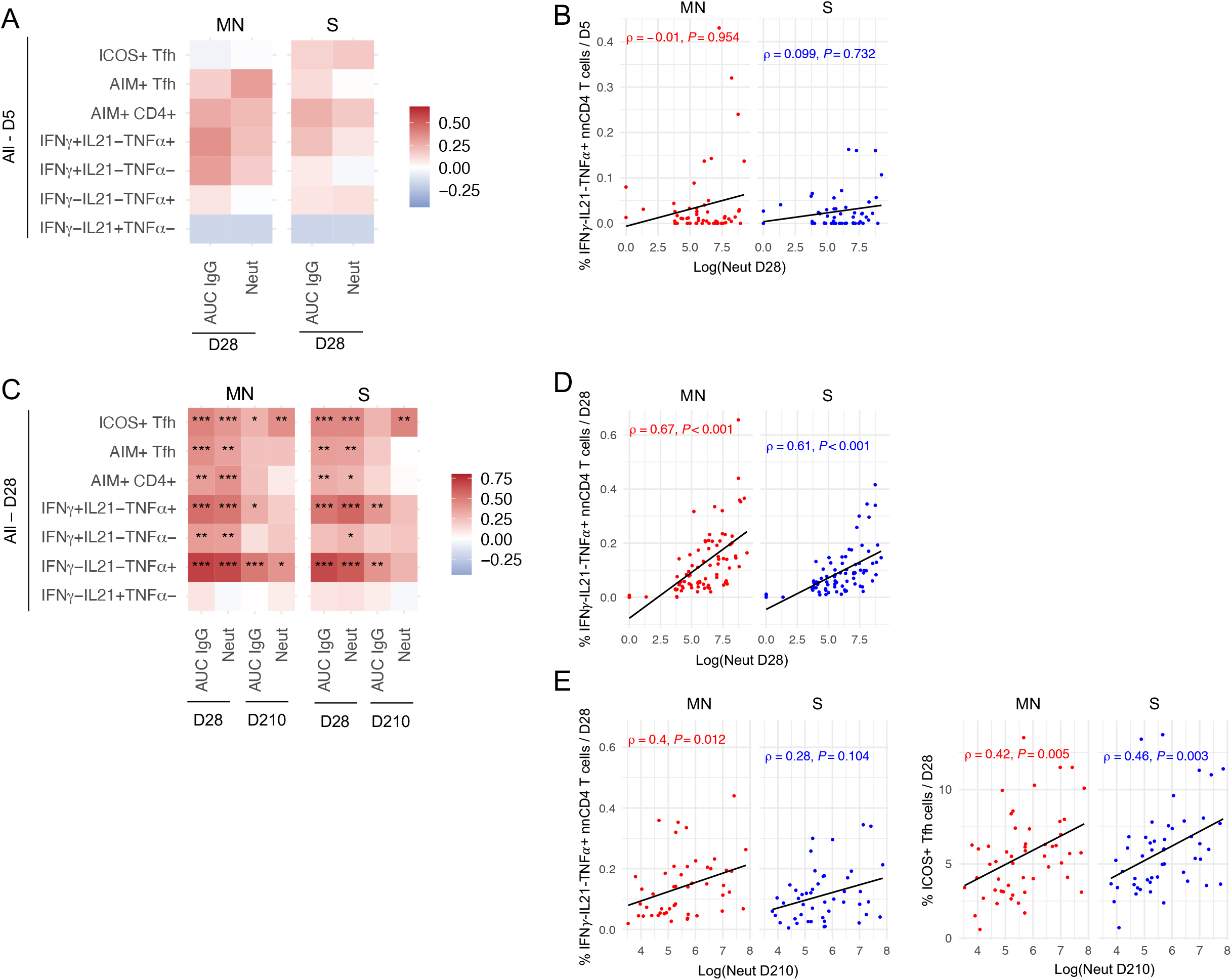
Late T cell responses are positively correlated with durability of antibody titers, while early T cell responses are not associated with the peak magnitude of antibody titers. A,C) In the heatmaps spearman’s correlations (Benjamini-Hochberg corrected) between indicated Tfh cell populations, AIM^+^ CD45RA^-^ CD4^+^ and cytokine^+^ non-naïve CD4^+^ T cell data, and antibody titers (S protein IgG binding (AUC IgG) or neutralizing antibody (Neut) are shown. The measurements that are significantly associated are indicated by asterisks (*=P<0.05, **=P<0.01, ***=P<0.001). A) Correlations are performed between indicated T cell data collected at D5 post-enrollment and indicated antibody titers at D28 post-enrollment. B) Scatter plots comparing MN protein-(left/red) and S protein-stimulated (right/blue) IFNγ^-^IL21^-^TNFα^+^-producing non-naïve CD4^+^ T cells collected on D5 (MN: n=55, S: n=51) post-enrollment with neutralizing antibody titers collected on D28 post-enrollment are depicted. C) Correlations are performed between indicated T cell data collected on D28 post-enrollment and indicated antibody titers at D28 or D210 post-enrollment. D) Scatter plots comparing antigen-stimulated IFNγ^-^IL21^-^TNFα^+^-producing non-naïve CD4^+^ T cells collected on D28 (MN: n=81, S: n=78) post-enrollment with neutralizing antibody titers collected on at D28 post-enrollment are shown. E) Scatter plots comparing antigen-stimulated IFNγ^-^IL21^-^TNFα^+^-producing non-naïve CD4^+^ T cells (left, MN: n=48, S: n=47) or ICOS^+^ cTfh cells (right, MN: n=52, S: n=50) collected on D28 with neutralizing antibody titers collected on D210 post-enrollment are shown. B,D,E) The neutralizing antibody titers are presented in natural logarithm and added +1 to allow for inclusion of participants with no neutralizing activity. The rho (ρ) and P values were calculated using spearman’s correlation (Benjamini-Hochberg corrected). The lines represent the fitted linear relationship between the indicated data.

We next examined whether SARS-CoV-2-specific CD4^+^ T cell responses measured later (D28) were associated with the durability of the antibody response, assessing correlations with antibody responses measured at D28 and month seven (D210). After multiple comparison correction, we found that SARS-CoV-2-specific CD4^+^ T cells that produce TNFα, in particular IFNγ^-^IL21^-^TNFα^+^cells, and activated ICOS^+^ cTfh cells on D28, correlated significantly and positively with both S binding IgG and neutralizing antibody titers measured on D28 and D210 (Figure 4C-E). Results were similar when analyzing the Lambda and placebo arm separately (Supplemental Figure 4G). In linear models adjusted for treatment arm, participant gender, and participant age, associations between MN protein-specific Log_10_ IFNγ^-^IL21^-^TNFα^+^-producing CD4^+^ T cells and antibody responses on D210 remained significant (Table 1). Similarly, associations between ICOS^+^ cTfh on D28 and antibody responses on D210 remained significant after multivariate adjustment (Table 1). Taken together, these findings show that SARS-CoV-2-specific IFNγ^-^IL21^-^TNFα^+^ CD4^+^ T cells and activated, ICOS^+^ cTfh on D28 correlate with SARS-CoV-2-specific antibody responses measured more than seven months post-infection.

**Table 1.**
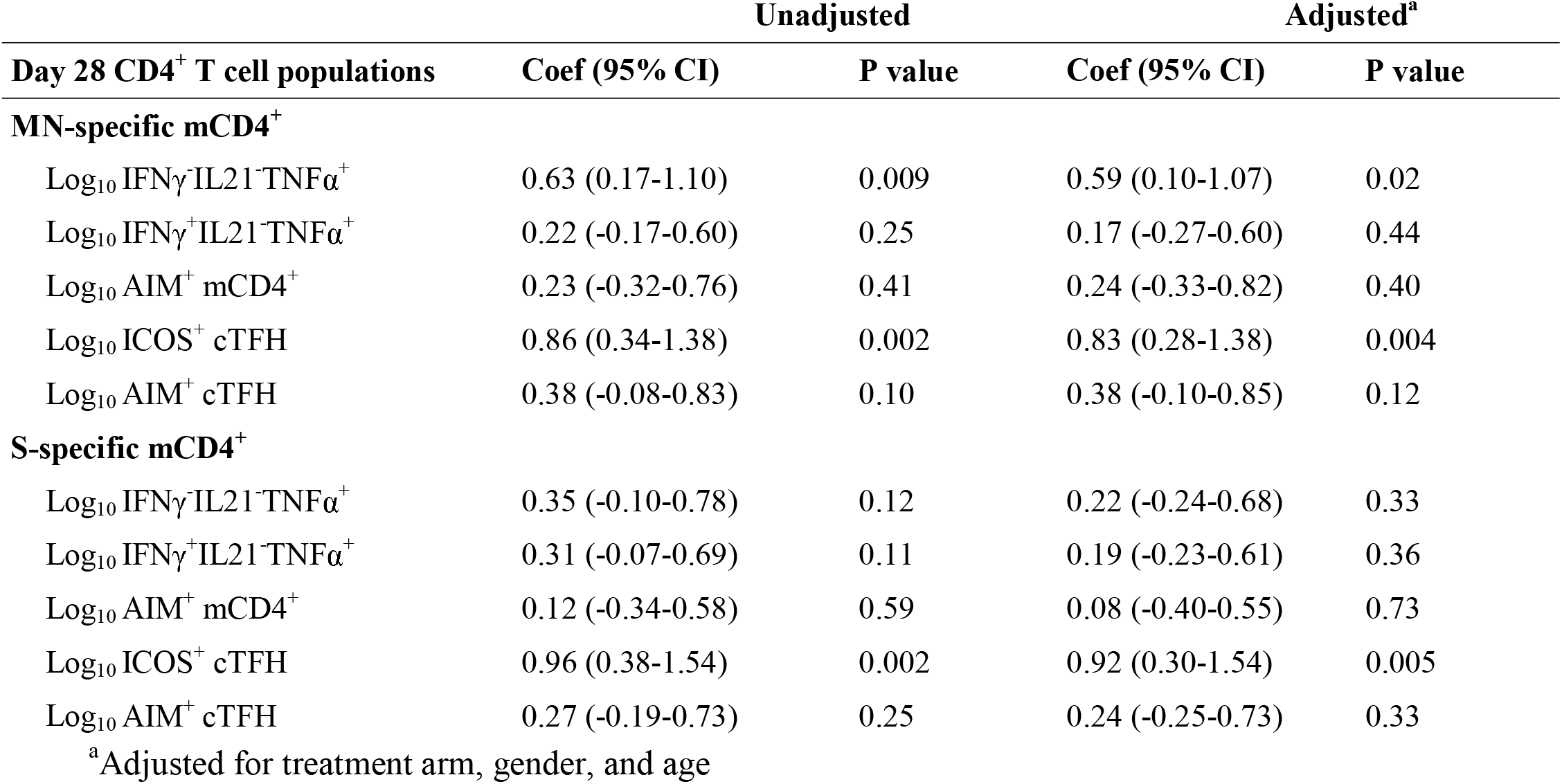
Linear model calculating associations between SARS-CoV-2-specific CD4^+^ T cell populations measured at D28 and antibody responses measured at D210

### Early proteomic and transcriptomic signatures associate with T cell responses on D28 post-enrollment

Given the robust associations between SARS-CoV-2-specific IFNγ^-^IL21^-^TNFα^+^ CD4^+^ T cells and ICOS^+^ cTfh, and durable neutralizing antibody titers, we next sought to identify early, infection-induced determinants of this response. We performed whole blood RNA sequencing and plasma proteomics by Olink on D0 and D5 post-enrollment (Hu et al., 2021), and correlated these measurements with the D28 SARS-CoV-2-specific CD4^+^ T cell response (Figure 5A). Several transcriptomic pathways associated with B cell activation, including B cell mediated immunity pathways, B cell receptor signaling, and immunoglobulin production, were associated with both SARS-CoV-2-specific CD4^+^ T cells producing IFNγ^-^IL21^-^TNFα^+^ and activated ICOS^+^ cTfh cells measured on D28 (Figure 5A, 5C). In addition, higher levels of several plasma proteins measured on D0 and D5 post-enrollment were associated with greater percentages of SARS-CoV-2-specific CD4^+^ T cell responses measured on D28, including IFNγ^-^IL21^-^TNFα^+^-producing cells (Figure 5A, Supplemental Table 2) and ICOS^+^ cTfh cells (Figure 5C and Supplemental Table 3). Several of these proteins were associated with both cellular populations, including monocyte-chemotactic protein 3 (MCP-3, also known as CCL7), eukaryotic translation initiation factor 4 gamma 1 (EIF4G1), DNA fragmentation factor subunit alpha (DFFA) and C-X-C motif chemokine ligand 1 (CXCL1) (Figure 5B, 5D). MCP-3 is a cytokine induced by IFNγ, and acts as a chemoattractant for activated leukocytes including CCR2^+^ monocytes (Menten et al., 1999; Proost et al., 1996). Although elevated levels of MCP3 have previously been implicated in the pathogenesis of severe COVID-19 (Hu et al., 2021; Merad & Martin, 2020; Yang et al., 2020), our data suggest that it may also be a useful early biomarker for the initiation of a robust, long-lived adaptive cellular immune response.

**Figure 5.**
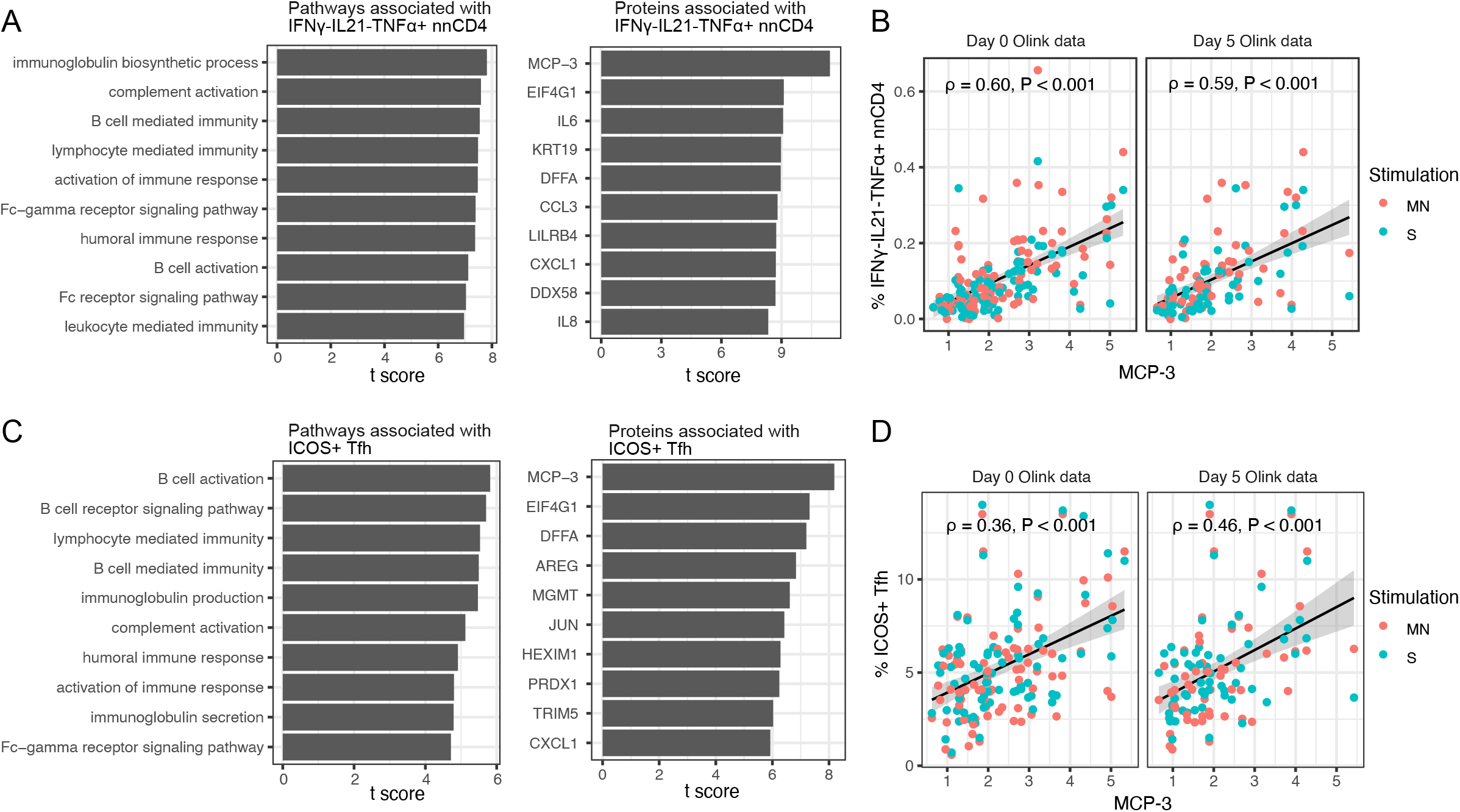
Associations between early immune response and T cell responses on D28 post-enrollment. Gene Ontology-based immune pathways (left) and plasma proteins (right) associated with the absolute percentage of SARS-CoV-2-specific IFNγ^-^IL21^-^TNFα^+^ non-naïve CD4^+^ T cells on D28. B) Scatter plots showing the correlation between the MCP-3 on D0 or D5 and the percentage of SARS-CoV-2-specific IFNγ^-^IL21^-^TNFα^+^ non-naïve CD4^+^ T cells on D28. C) Gene Ontology-based immune pathways (left) and plasma proteins (right) associated with the absolute percentage of ICOS^+^ cTfh cells on D28. D) Scatter plots showing the correlation between the MCP-3 on D0 or D5 and ICOS^+^ cTfh cells on D28. B,D) The color of the points represents the stimulating antigens. We used regression models to test the association while controlling for the sampling time of immune pathways or proteins (D0 or D5) and the stimulation type (MN or S) of the T cell response.

### SARS-CoV-2-specific CD4^+^ T cells producing IFNγ and TNFα and AIM^+^ cTfh cells are boosted following COVID-19 mRNA vaccination

COVID-19 vaccines elicit robust humoral and CD4^+^ T cell responses among participants who had not been previously exposed to SARS-CoV-2 (Mateus et al., 2021; Mulligan et al., 2020; Sahin et al., 2020; Woldemeskel et al., 2021). However, little has been reported to date on the CD4^+^ T cell response following mRNA vaccination among individuals who have experienced SARS-COV-2 infection (Goel et al., 2021; Painter et al., 2021). We evaluated the SARS-CoV-2-specific CD4^+^ T cell response in study participants who received two doses of a COVID-19 mRNA vaccine either prior to the month seven (n=2) or month ten (M10) (n=18) follow-up visits, and compared these changes to unvaccinated participants. As expected, we found that the magnitude of S protein-specific, but not MN protein-specific, IFNγ^+^IL21^-^TNFα^-^-and IFNγ^+^IL21^-^TNFα^+^-producing CD4^+^ T cells was increased in the post-vaccination samples only (Figure 6A and Supplemental Figure 7A). The M10/D28 ratio of S protein-specific IFNγ^+^IL21^-^TNFα^-^ cells was 3.3-fold higher, and IFNγ^+^IL21^-^TNFα^+^ cells 4.9-fold higher, comparing vaccinated to unvaccinated participants. In contrast, we observed a more modest, 1.4-fold increase in the M10/D28 ratio of S protein-specific IFNγ^-^IL21^-^TNFα^+^ cells (Figure 6B). These differences did not appear to be driven by time since second dose (Supplemental Figure 7B). Additionally, vaccinated individuals had a 4.3-fold higher M10/D28 ratio of S protein-specific AIM^+^ cTfh cells compared to unvaccinated participants (Figure 6C), without boosting of AIM^+^ CD4^+^ T cells or activated cTfh cells (Supplemental Figure 7C, 7D). Together, these findings show that mRNA vaccination boosted S protein-specific CD4^+^ T cells that produced IFNγ, with or without TNFα, along with AIM^+^ cTfh cells.

**Figure 6.**
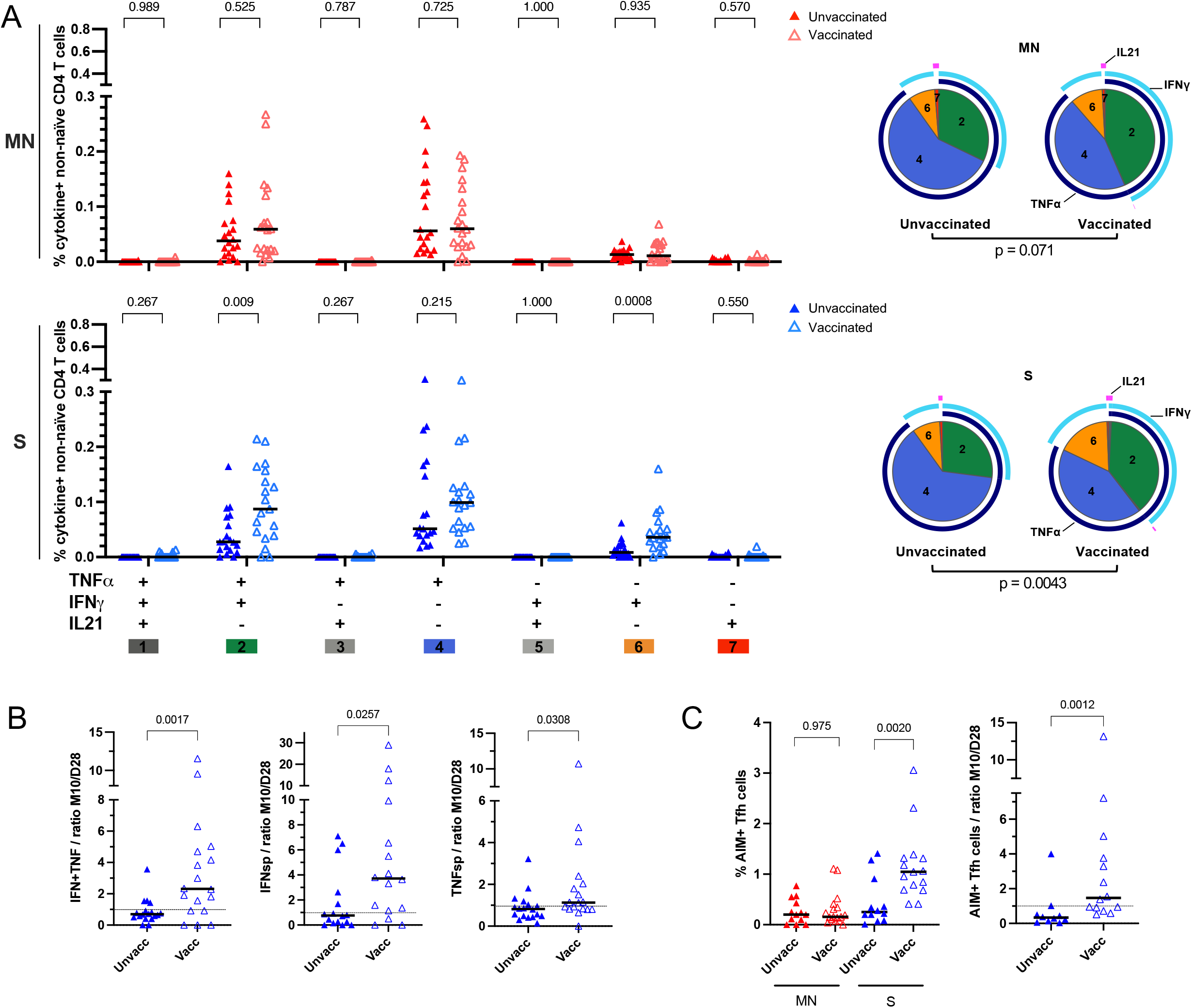
mRNA vaccination boosts IFNγ^+^IL21^-^TNFα^-^-and IFNγ^+^IL21^-^TNFα^+^-producing CD4^+^ T cells and AIM^+^ cTfh cells. PBMCs of COVID-19 outpatients on month ten (M10) post-enrollment were stimulated with S or MN proteins in vitro, stained and analyzed by flow cytometry. A) The absolute percentage (scatter plot, black line = median) of each individual combination of TNFα, IFNγ and IL21^-^producing non-naïve CD4^+^ T cells stratified by vaccination status (closed triangle: unvaccinated n=20, open triangle: vaccinated, n=20). Top panel depicts MN protein-stimulated CD4^+^ T cells (red) and bottom panel S protein-stimulated CD4^+^ T cells (blue). P values shown above the scatter plots are calculated using the Mann Whitney test. The pie charts represent the relative proportion (mean) of each individual combination of cytokine-producing non-naïve CD4^+^ T cells are shown stratified by antigenic stimuli. The P value shown under the pie charts is calculated using a partial permutation test. B) The ratio between M10 and D28 of the absolute percentage of IFNγ^+^IL21^-^TNFα^+^- (IFN+TNF, left), IFNγ^+^IL21^-^TNFα^-^-(IFNsp, middle) or IFNγ^-^ IL21^-^TNFα^+^-producing (TNFsp, right) non-naïve CD4^+^ T cells stimulated with the S protein is depicted (scatter plot, black line = median). C) The absolute percentage (left) and the ratio between M10 and D28 of AIM^+^ (CD137^+^/OX40^+^) cTfh cells (right) stimulated with the MN (red) or S proteins (blue) and stratified by vaccination status are shown. B, C) The P values shown are calculated using the Mann Whitney test.

## Discussion

In this longitudinal study of SARS-CoV-2-specific CD4^+^ T cell magnitude and quality in outpatients enrolled in a Phase 2 clinical trial of Peginterferon Lambda, we identified a shift in the quality of the SARS-CoV-2-specific CD4^+^ T cell response from IFNγ^-^producing at early timepoints to TNFα-producing response at later timepoints (≥D28). IFNγ^-^IL21^-^TNFα^+^ CD4^+^ T cells were the predominant cytokine-producing T cell population following infection and sustained up to month ten post-enrollment. Percentages of TNFα-producing CD4^+^ T cells were positively correlated with AIM^+^ T cell populations and activated ICOS^+^ cTfh cells. Higher percentages of TNFα-producing CD4^+^ T cells and ICOS^+^ cTfh cells were more commonly found in participants who had experienced a more severe infection and were positively associated with higher levels of the chemokine CCL7/MCP-3 at the time of enrollment. Percentages of IFNγ^-^ IL21^-^TNFα^+^CD4^+^ T cells, along with activated ICOS^+^ cTfh cells, were positively correlated with neutralizing antibodies measured up to seven months post-enrollment. These cells were only modestly boosted following mRNA vaccination in comparison to other cellular populations, including S protein-specific AIM^+^ cTfh cells and IFNγ^+^-producing CD4^+^ T cells.

Polyfunctional CD4^+^ Th1 cells producing IFNγ and TNFα (along with IL2) have been thought to be important in response to some viral infections, including HIV and influenza (Betts et al., 2006; Lin et al., 2015; Seder et al., 2008). This polyfunctional Th1 cytokine profile has been observed among convalescent mild and severe COVID-19 patient in several studies (Braun et al., 2020; Breton et al., 2021; Grifoni et al., 2020; Neidleman et al., 2020; Peng et al., 2020; Rydyznski Moderbacher et al., 2020; Sekine et al., 2020; Weiskopf et al., 2020; Zuo et al., 2021), although the importance of this versus other cytokine-producing populations of circulating SARS-CoV-2-specific CD4^+^ T cells remains unclear. However, many of these studies were cross-sectional in nature, with significant heterogeneity observed (Rydyznski Moderbacher et al., 2020; Sekine et al., 2020; Tan et al., 2021; Weiskopf et al., 2020; Zhou et al., 2020). In the present study, we detected a substantial proportion of CD4^+^ T cells producing both IFNγ and TNFα (along with IL2) that peaked in percentage early in convalescence. In contrast, at later timepoints, TNFα-producing cells, without IFNγ, were the predominant population detected in our ICS assay. This finding is particularly relevant given the development of clinical assays designed to detect SARS-CoV-2-specific T cell responses via measurement of IFNγ release (Murugesan et al., 2021), and suggest that assays measuring SARS-CoV-2-specific TNFα production could be considered as a tool to define previous exposure to SARS-CoV-2.

In addition to measuring SARS-CoV-2-specific T cells by ICS, we also utilized an AIM assay employing markers for both CD4^+^ T cells and cTfh cells, and similar with published reports (Dan et al., 2021; Jung et al., 2021), detected SARS-CoV-2-specific AIM^+^ CD4^+^ T cells up to ten months post-enrollment. Moreover, we observed that cytokine-producing CD4^+^ T cell populations -in particular, TNFα single positive cells - positively and significantly correlated with AIM^+^ CD4^+^ T cells, activated ICOS^+^ cTfh cells, and AIM^+^ cTfh cells. Given their central-memory like phenotype, we speculate that SARS-CoV-2-specific TNFα single positive cells may represent a memory pool of either memory Th1 or Tfh cells (Lönnberg et al., 2017) or stem-cell like memory cells (Jung et al., 2021); molecular work is ongoing to identify transcription factors that might help elucidate the ontogeny of these cells. Our observations that several plasma proteins, including the chemokine MCP-3/CCL7, measured at the time of infection strongly correlate with higher percentages of these cells on D28 suggests that a robust innate immune response at the time of infection may lead to greater expansion of this cell subset; however, this immune activation may also represent a double-edged sword given its association with early disease progression (Hu et al., 2021; Merad & Martin, 2020; Yang et al., 2020). No correlation was observed between IL21^-^producing CD4^+^ T cells and cTfh cells. This was unexpected given that IL21 is thought to be a canonical cTfh cytokine (Crotty, 2019; Eto et al., 2011; Hale & Ahmed, 2015; Rasheed et al., 2013). Furthermore, we were unable to detect any antigen-specific IL10 production. One possible explanation for these findings is that our ICS assay was not optimized to detect SARS-CoV-2-specific IL21 or IL10 production, although we and others have successfully used a similar approach to detect these CD4^+^ T cell cytokines in other settings (Jagannathan et al., 2014). Alternatively, it is possible that cTfh are not representative of germinal center Tfh in this setting, or that these cytokines may not be induced in the SARS-CoV-2-specific CD4^+^ T cell compartment following infection, although this will require further validation in other cohorts and settings.

Although several studies have reported correlations between SARS-CoV-2-specific CD4^+^ T cell responses and SARS-CoV-2-specific antibody titers measured at either the peak of response or at concurrent timepoints (Boppana et al., 2021; Dan et al., 2021; Ni et al., 2020; Peng et al., 2020; Rydyznski Moderbacher et al., 2020; Zuo et al., 2021), there is limited data correlating infection-induced CD4^+^ T cell responses with antibodies measured prospectively. Although SARS-CoV-2-specific, polyfunctional IFNγ and TNFα co-producing CD4^+^ T cells at D5 correlated with the neutralizing antibody response at D28 by Spearman correlation, this finding was not significant after multiple comparison correction. In contrast, we observed that TNFα-single positive cells measured on D28, rather than polyfunctional, IFNγ and TNFα co-producing cells, were most strongly correlated with neutralizing antibodies measured seven months post-enrollment. We also observed a significant positive correlation between activated, ICOS^+^ cTfh cell percentages on D28 and neutralization antibodies on month seven. This suggests that SARS-CoV-2-specific TNFα-producing CD4^+^ T cells and activated cTfh cells measured weeks after infection may serve as a useful correlate to identify individuals who exhibit durable antibodies. Furthermore, additional characterization of these cellular populations would help identify strategies to determine whether boosting of this response could increase durability of neutralizing antibodies.

Given our study design, we were uniquely positioned to compare CD4^+^ T cell responses among previously infected study participants who did and did not receive two doses of a COVID-19 mRNA vaccine during follow-up. We found that mRNA vaccination particularly boosts S protein-specific CD4^+^ T cells that produce IFNγ, with or without TNFα, along with AIM^+^ cTfh cells. There was only modest boosting of TNFα single positive cells, and no boosting of MN protein–specific cells. These findings are consistent with a recent systems analysis of immune responses following the BNT162b2 mRNA vaccine (Arunachalam et al., 2021). In that study, Arunachalam and colleagues found that vaccination resulted in robust expansion of IFNγ^-^producing CD4^+^ T cells, although they observed no significant correlation between levels of IFNγ producing CD4^+^ T cells and the neutralizing antibody response measured prospectively. In another study by Painter and colleagues, the percentage of antigen-specific cTfh cells prior to the second dose of mRNA vaccination correlated with post-second dose neutralizing antibody titers. In contrast, pre-second dose antigen-specific Th1 cells were less well correlated, suggesting distinct associations between Th populations and vaccine-elicited immune responses (Painter et al., 2021). Together, these data suggest that the quality of the CD4^+^ T cell response differs between natural infection and vaccination, although the downstream impact of different CD4^+^ T cell responses on the neutralizing antibody response and protective immunity remain unclear.

This study had some limitations. Half of the study participants in this trial received an investigational Type III IFN at the time of infection. However, in this study, this agent neither shortened the duration of viral shedding nor symptoms (Jagannathan et al., 2021), nor did we observe any significant impact on either innate (Hu et al., 2021) or adaptive (Chakraborty et al., 2021) immune responses between arms, allowing us to utilize data from both arms to improve statistical power. In addition, stratified analyses, and multivariate models adjusted for treatment arm as a covariate, confirmed pooled results. Although we would have liked to have assessed for correlations with protection against reinfection, only one participant had evidence of reinfection during follow-up. Future larger cohorts, and/or case/control designs, will be required to address this question. Finally, we only utilized data assessing responses to the original consensus strain, and do not present data on responses to variants. However, others have recently reported that infection-induced SARS-CoV-2 T cell responses have broad reactivity against viral variant proteins (Keeton et al., 2021; Tarke et al., 2021).

By providing insight into the shifting quality of the SARS-CoV-2-specific CD4^+^ T cell response following infection, how this is impacted by vaccination, and which features most strongly correlate with immune effector mechanisms, our study adds to our growing understanding of the memory T cell response to SARS-CoV-2. Given ongoing SARS-CoV-2 transmission and increasing risk of both re-infections and breakthrough infections following vaccination, identification of correlates of protective immunity remains critical in the design of next generation COVID-19 vaccination strategies.

## Data Availability

All data produced in the present study are available upon reasonable request to the authors.

https://www.ncbi.nlm.nih.gov/geo/query/acc.cgi?acc=GSE178967

## Acknowledgments

This study was supported by NIH/NIAID (U01 AI150741-01S1 to Z.H., S.T., I.R.B., B.G., T.T.W., and P.J.), the Stanford’s Innovative Medicines Accelerator, and a Fastgrant (C.A.B.). The Lambda clinical trial was funded by anonymous donors to Stanford University, and Peginterferon Lambda provided by Eiger BioPharmaceuticals. The funders had no role in data collection and analysis or the decision to publish.

We thank all study participants who participated in this study, the study team for their tireless work, and Thanmayi Ranganath, Nancy Q. Zhao, Aaron J. Wilk, Rosemary Vergara, Julia L. McKechnie, Giovanny J. Martínez-Colón, Arjun Rustagi, Geoff Ivison, Ruoxi Pi, Madeline J. Lee, Taylor Hollis, Georgie Nahass, Kazim Haider, and Laura Simpson for assistance with processing samples. We also thank our colleagues at Stanford University Occupational Health and at San Mateo Medical Center for participant referrals. The Stanford REDCap platform (http://redcap.stanford.edu) is developed and operated by Stanford Medicine Research IT team. The REDCap platform services at Stanford are subsidized by a) Stanford School of Medicine Research Office, and b) the National Center for Research Resources and the National Center for Advancing Translational Sciences, National Institutes of Health, through grant UL1 TR001085.

## Author Contributions

Conceptualization, K.v.d.P., A.S.K., S.C., Z.H., B.J.S., J.P., J.R.A., U.P., T.T.W. and P.J.; Methodology, K.v.d.P., A.S.K., S.C., Z.H., B.L.S., J.R.A, G.S.T., C.A.B., S.T., I.R.B., B.G., T.T.W., P.J.; Formal Analysis, K.v.d.P., A.S.K., S.C., Z.H., P.J.; Investigation, K.v.d.P., D.A.M.M., S.C., B.L.S., K.B.J., H.B., J.P., J.R.A., K.D.P., M.C.T., D.R.R.B., L.d.l.P., G.S.T., C.A.B., U.P., T.T.W., P.J.; Data Curation, K.v.d.P., D.A.M.M., S.C., B.L.S., K.B.J., K.D.P., M.C.T., D.R.R.B., L.d.l.P., C.A.B., U.P., T.T.W., P.J.; Visualization, K.v.d.P.; Supervision, P.J.; Project Administration, D.A.M.M., P.J.; Funding Acquisition, C.A.B, B.G., U.P., T.T.W., P.J.; Writing – Original Draft, K.v.d.P., P.J.; Writing – Review & Editing, all authors.

## Declaration of interests

The authors declare no competing interests.

## Methods

### Clinical cohort and samples

The samples used were from 109 participants enrolled in a Phase 2, single-blind, randomized placebo-controlled trial evaluating the efficacy of Peginterferon Lambda-1a in SARS-CoV-2 infected outpatients (Jagannathan et al., 2021). The trial was registered at ClinicalTrials.gov (NCT04331899) and was performed as an investigator-initiated clinical trial with the FDA (IND 419217). In brief, symptomatic outpatients aged 18-71 who tested positive for reverse transcription-polymerase chain reaction (RT-PCR) detection of SARS-CoV-2 were eligible to participate in the study barring any signs of respiratory distress. Asymptomatic patients were eligible if infection was the initial diagnosis of SARS-CoV-2 infection. Full eligibility and exclusion criteria are provided in the study protocol and have been published (Jagannathan et al., 2021). On the day of enrollment, participants were randomized 1:1 to receive a single subcutaneous injection of Lambda or saline placebo. For the clinical trial, participants were followed for 28 days with an at home daily symptom survey (REDCap Cloud) and daily in-home assessments of temperature and oxygen saturation. In-person follow-up visits were conducted on day 1, 3, 5, 7, 10, 14, 21, 28 for symptom assessments, collection of oropharyngeal swabs, safety labs (day 5, 14), and peripheral blood biobanking (day 5, 14, 28.) Participant baseline demographics and clinical characteristics are depicted in Supplemental Table 1. Participants were invited to return for long-term follow-up visits at month four, month seven and month ten post-enrollment for a symptom survey, clinical assessment, assessment of vaccination status, and peripheral blood biobanking. Overall, 20 participants received 2 doses of COVID-19 mRNA vaccine before month seven (n=2) or month ten (n=18) follow-up visits (median 30 days since second dose at time of sampling, range 6-78 days). We also collected blood and serum samples from healthy volunteers, without known SARS-CoV-2 exposure, aged 22-35 to serve as our non-exposed controls. All participants gave written informed consent. The study protocol used was approved by the Institutional Review Board of Stanford University.

### PBMC/Serum Isolation

Blood was collected from study participants in two BD Vacutainer® CPT™ Mononuclear Cell Preparation Tubes, which provide a fully-closed system for separation of mononuclear cells from whole blood. The tubes were centrifuged at 1800g for 20 minutes, following centrifugation plasma was separated into a 15mL centrifuge tube, centrifuged again at 1200g for 10 minutes, then aliquot and stored at −80°C. The PBMC layer was transferred to a 50mL centrifuge tube, washed twice with Gibco DPBS and centrifuged at 300g for 10 minutes. After counting, the cells were aliquoted in 90% Fetal Bovine Serum and 10% DMSO at 5 million cells per vial. PBMC vials were stored overnight at −80°C, then transferred to liquid nitrogen storage. Whole blood was collected in Paxgene Tubes.

### Peptides

For *in vitro* stimulation experiments, we used PepTivator® SARS-CoV-2 lyophilized peptide pools from Miltenyi Biotec. We used Prot_M and Prot_N, consisting of complete sequences of the membrane ‘M’ glycoprotein (GenBank MN908947.3, Protein QHD43419.1) and nucleocapsid ‘N’ phosphoprotein (GenBank MN908947.3, Protein QHD43423.2) of SARS-CoV-2. For the spike ‘S’ glycoprotein of SARS-CoV-2 we used the combination of Prot_S1 and Prot_S peptide pools, which covers the aa sequence 1-692 of the N-terminal S1 domain (GenBank MN908947.3, Protein QHD43416.1) and the immunodominant sequence domains, within aa 304-1273, of the S protein (GenBank MN908947.3, Protein QHD43416.1), respectively.

### Functional T cell assays

#### Intracellular cytokine staining

After thawing, PBMCs were rested overnight in complete RPMI (RPMI (Corning) supplemented with 10% FBS (Gibco), 100 IU Penicillin (Corning), 100 ug/ml Streptomycin (Corning), 1 mM Hepes (Corning) and 2 mM L-glutamine (Corning)). We prepared 96-well U-bottom plates with 1×10^6 cells in 200uL of complete RPMI and then rested overnight at 37°C in a CO2 incubator. Cells were cultured in presence of either SARS-CoV-2 peptides (1 ug/ml), PMA (300 ng/ml) and Ionomycin (1.5 ug/ml) as positive control, or media as a negative control for 6 hours at 37°C the following morning. Brefeldin-A (BD Pharmingen), Monensin (BD Pharmingen) and CD107a were present in all conditions from the start of incubation. Thereafter, cells were washed and surface stained for CCR7 for 15 min at 37°C, which was followed by the remaining surface stain for 30 min at room temperature (RT) in the dark. Afterwards, cells were washed twice with PBS containing 0.5% BSA and 2 mM EDTA, then fixed/permeabilized (FIX & PERM® Cell Permeabilization Kit, Invitrogen) and stained with intracellular antibodies for 20 min at RT in the dark. A complete list of antibodies is listed in Supplemental Table 4.

#### Activation Induced Marker (AIM) Assay

Thawed PBMCs were prepared in 96-well U-bottom plates at 1×10^6 cells, in 200uL of complete RPMI and rested for 1 hour at 37°C. The cells were stimulated overnight in presence of either SARS-CoV-2 peptides (1 ug/ml), PHA (2 ug/mL) as positive control, or media as a negative control at 37°C. Thereafter, cells were washed and surface stained for CXCR5 for 30 min at 37°C. This was followed by the remaining surface stain for 30 min at RT in the dark. Antibodies used are listed in Supplemental Table 5. All samples were analyzed on an Attune NXT flow cytometer and analyzed with FlowJo X (Tree Star) software.

### ELISA and neutralizing assay

ELISA and neutralizing assay were performed as described in detail previously (Chakraborty et al., 2021). IgG antibody titers against the SARS-CoV-2 spike receptor binding domain (RBD) were assessed at enrollment, day 28 and 210 post-enrollment by ELISA. Briefly, 96 Well Half-Area microplates (Corning (Millipore Sigma)) plates were coated with antigens at 2 μg/mL in PBS for 1 hour at RT. Next, the plates were blocked and plasma was diluted fivefold starting at 1:50, before adding 25μL of the diluted plasma to each well and incubated for 2 hours at RT. This is followed by adding 25 μL of 1:5000 diluted horse radish peroxidase (HRP) conjugated anti-Human IgG secondary antibodies (Southern Biotech) and incubated for 1 hour at RT. The plates were developed and absorbance was measured at 450nm (iD5 SPECTRAmax, Molecular Devices). All data were normalized between the same positive and negative controls and the binding area under the curve (AUC) reported.

The generation of vesicular stomatitis virus (VSV) pseudotyped with the S of SARS-CoV-2 were made as described previously (Chakraborty et al., 2021). In short, neutralization assays were performed by seeding Vero cells (ATCC CCL-81) 24 hours prior to the assay and by plating patient plasma on a seperate plate in serially 5-fold dilution, which was followed by adding 25 μL of SARS-CoV-2 pseudo-typed VSV particles to the wells on the dilution plate and incubated at 37°C for 1 hour. Prior to infection, Vero cells were washed twice with 1X PBS and then 50 μL of the incubated pseudo-typed particles and patient plasma mixture was transferred onto the Vero cells, followed by a 24 hour incubation at 37°C and 5% CO2. Afterwards, viral infection was analyzed by quantifying the number of GFP-expressing cells using a Celigo Image Cytometer. First, the percent infection was calculated based on the ‘virus only’ controls and then the percent inhibition was determined by subtracting the percent infection from 100. A non-linear curve and the half-maximal neutralization titer (NT50) were generated using GraphPad Prism.

### Whole blood transcriptomics and data analysis

Novogene Corporation, Inc. executed whole blood transcriptomics. In brief, using whole blood samples collected in Paxgene Tubes and treated with Proteinase K, RNA extraction was done using Quick-RNA MagBead Kit (R2132) on KingFisher. Thereafter, quality control checks were performed using a Qubit and Bioanalyzer 2100. Libraries were prepared using ZymoSeq RiboFree Total RNA Library Kit (R3000) and sequencing took place on a Nova6000 on an S4 lane, 30M paired reads, PE 150. Data analysis has been described in detail by Hu et al (Hu et al., 2021).

### Plasma proteomics using Olink panels

Proteins in plasma were measured using Olink multiplex proximity extension assay (PEA) inflammation panel and immune response panel (Olink proteomics, www.olink.com) according to the manufacturer’s instructions (Hu et al., 2021).

### Statistical analysis

All statistical analyses were performed using Prism 6.0 (GraphPad), STATA version 16 (College Station), SPICE v.6 (NIAID) (Roederer et al., 2011), and/or R. Percentages of SARS-CoV-2-specific cytokine producing T cells (alone or in combination) are reported after background subtraction of the percentage of the identically gated population of cells from the same sample stimulated with media control. Background-subtracted responses were considered positive if >0.01% parent population. Samples included in experiments with poor or no viability (determined by Live/Dead Aqua stain (Invitrogen)) were excluded during analysis. Age categories were defined by participant age quartiles. Comparisons of cytokine percentages between groups were done using the Mann Whitney test. The Wilcoxon matched-pairs signed-rank test or Friedman test with Dunn’s multiple comparison test were used to compare paired data. Statistical analyses of global cytokine profiles (pie charts) were performed by partial permutation tests using SPICE. Continuous variables were compared using Spearman correlation with Benjamini-Hochberg multiple comparison correction. For multivariate linear regression models, non-normal variables were log-transformed. Linear regression was used to estimate associations between T cell responses measured on day 28 and neutralizing antibody levels measured on month four, adjusting for participant sex, treatment arm, and age.

We tested associations between plasma proteins measured by Olink, whole blood transcriptomics, and CD4^+^ T cell responses using regression models and the lm function in R, adjusting for time since symptom onset and the stimulation type (MN or S) of the T cell response (T cell responses ∼ measurements + Stim + time + time^2^). Both the first and second order term of time were included in the model to adjust for the non-linear relationship between time and immune response, as described in an early publication (Hu et al., 2021). The P value of the regression coefficient of the measurement term is used to determine the significance of association between T cell response and the immune measurements. The False Discovery Rate (FDR) method is used to adjust for the multiple hypothesis testings when assessing the association between T cell response and the immune measures (plasma proteins and immune pathways). Two-sided p-values were calculated for all test statistics and P<0.05 was considered significant.

## Supplemental information

**Supplemental Information PDF**

**Supplemental Table 2. Plasma proteins significantly associated with IFNγ^-^IL21^-^TNFα^+^ SARS-CoV-2-specific T cells**

**Supplemental Table 3. Plasma proteins significantly associated with ICOS^+^ cTfh cells**

## Notes

### Competing Interest Statement

The authors have declared no competing interest.

### Clinical Trial

NCT04331899; IND 419217

### Funding Statement

This study was funded by NIH/NIAID (U01 AI150741-01S1, UL1 TR001085), the Stanfords Innovative Medicines Accelerator (Chem-H IMA), and a Fastgrant.

### Author Declarations

Study was approved by Institutional Review Board of Stanford University (protocol 57230)

