## Supplemental data for "TNFα-producing CD4^+^ T cells dominate the SARS-CoV-2-specific T cell response in COVID-19 outpatients and are associated with durable antibodies"

**Supplemental Table 1. Baseline Characteristics**

| | Treatment arm | | |
|---|---|---|---|
| | **Lambda (N=55)** | **Placebo (N=54)** | **Overall (N=109)** |
| **Age in years**, median (range) | 38 (21-66) | 35 (20-71) | 37 (20-71) |
| **Male**, n (%) | 33 (60.0%) | 31 (57.4%) | 64 (58.7%) |
| **Race / Ethnicity**, n (%) | | | |
| Latinx | 32 (58.2%) | 37 (68.0%) | 67 (62.0%) |
| White | 18 (30.0%) | 12 (25.0%) | 30 (27.8%) |
| Asian | 4 (5.0%) | 5 (6.7%) | 9 (8.3%) |
| Native Hawaiian or other Pacific Islander | 2 (3.3%) | 0 (0%) | 2 (1.9%) |
| **BMI (kg/m$^2$)**, median (IQR) | 27.6 (25.4-31.1) | 28.5 (24.8-32.3) | 27.7 (24.9-32.0) |
| **Comorbid conditions** | | | |
| Hypertension | 9 (15.0) | 5 (8.3) | 14 (11.7) |
| Diabetes | 4 (6.7) | 8 (13.3) | 12 (10.0) |
| Asthma | 2 (3.3) | 2 (3.3) | 4 (3.3) |
| Heart Disease | 3 (5.0) | 1 (1.7) | 4 (3.3) |
| **Asymptomatic at baseline**, n (%) | 6 (10.0%) | 2 (3.3%) | 8 (6.7%) |
| **Duration of symptoms in days prior to randomization**, median (IQR) [a] | 4 (3-6) | 5 (3-5) | 5 (3-6) |
| **Baseline laboratory values**, median (IQR) | | | |
| White blood cell (WBC) count, cells/µl | 5.5 (4.3-6.8) | 5.6 (4.0-7.5) | 5.5 (4.1-7.1) |
| Absolute lymphocyte count (ALC), cells/µl | 1.5 (1.2-1.9) | 1.5 (1.2-2.3) | 1.5 (1.2-2.2) |
| Aspartate aminotransferase, IU/L | 31 (26-41) | 30 (25-39.3) | 30 (25-41) |
| Alanine aminotransferase, IU/L | 32.5 (21-52.3) | 30.5 (23-47.5) | 31.5 (22-50.3) |
| **Baseline oropharyngeal SARS-CoV-2 cycle threshold, median (IQR)** [b] | 30.9 (26.4-33.8) | 29.3 (26.4-34.3) | 30.3 (26.4-34.3) |
| **Baseline SARS-CoV-2 IgG seropositivity**, n (%) | 17 (31.5%) | 24 (44.4%) | 41 (38.0%) |

IQR inner quartile range
[a]Among n = 103 participant who reported symptoms prior to randomization (n = 48 in lambda and n = 55 in placebo)
[b]Among n = 87 participants with detectable OP virus (n = 44 in lambda and n = 43 in placebo)

#### Supplemental Table 4: ICS antibody panel

| **ICS Antibody Panel** | | | | | |
|---|---|---|---|---|---|
| **Surface Antibodies** | **Fluorochrome** | **Clone** | **Vendor** | **Catalog** | **Amount Per 50 uL** |
| CCR7 | BV421 | G043H7 | BioLegend | 353208 | 2.5 uL |
| CD14<br>CD19<br>LIVE/DEAD | BV510<br>BV510<br>Aqua | <br>M5E2<br>HB19 | BioLegend<br>BioLegend<br>Invitrogen | 301842<br>302242<br>L34965 | 0.5 uL<br>0.5 uL<br>0.25 uL |
| CD45RA | BV605 | HI100 | BioLegend | 304134 | 0.4 uL |
| CD4 | BV650 | RPA-T4 | BioLegend | 300536 | 1 uL |
| CD8A | BV785 | RPA-T8 | BioLegend | 301046 | 1 uL |
| CD107A | FITC | H4A3 | BioLegend | 328606 | 1 uL |
| CD3 | APC-H7 | SK7 | BD Biosciences | 560176 | 2.5 uL |
| **Intracellular Antibodies** | **Fluorochrome** | **Clone** | **Vendor** | **Catalog** | **Amount Per 50 uL** |
| IFNγ | PerCP Cy5.5 | 4S.B3 | BioLegend | 502526 | 0.5 uL |
| IL21 | eFluor660 | eBio3A3-N2 | eBioscience | 50-7219-42 | 1.25 uL |
| TNF | AF700 | MAb11 | BD Biosciences | 557996 | 0.5 uL |
| IL2 | PE | MQ1-17H12 | BioLegend | 500307 | 2 uL |
| IL10 | PE | JES3-19F1 | BD Biosciences | 559330 | 2 uL |

#### Supplemental Table 5: AIM antibody panel

| AIM Antibody Panel | | | | | |
|---|---|---|---|---|---|
| Antibody | Fluorochrome | Clone | Vendor | Catalog | Amount Per 50 uL |
| PD-1 | BV421 | EH12.2H7 | BioLegend | 329920 | 2 uL |
| CD14 | BV510 | | BioLegend | 301842 | 0.5 uL |
| CD19 | BV510 | M5E2 | BioLegend | 302242 | 0.5 uL |
| LIVE/DEAD | Aqua | HB19 | Invitrogen | L34965 | 0.25 uL |
| CD45RA | BV605 | HI100 | BioLegend | 304134 | 0.4 uL |
| CD4 | BV650 | RPA-T4 | BioLegend | 300536 | 1 uL |
| CXCR5 | BV711 | J252D4 | BioLegend | 356934 | 1 uL |
| CD8A | BV785 | RPA-T8 | BioLegend | 301046 | 1 uL |
| CD69 | FITC | FN50 | BD Biosciences | 555530 | 1.25 uL |
| OX40 | PE | Ber-ACT35 | BioLegend | 350004 | 2.5 uL |
| CD137 | APC | 4B4-1 | BioLegend | 309810 | 2.5 uL |
| CD3 | AF700 | SK7 | BioLegend | 344822 | 1 uL |
| ICOS | APC-Cy7 | C398.4A | BioLegend | 313530 | 0.8 uL |

#### Supplemental Figure 1

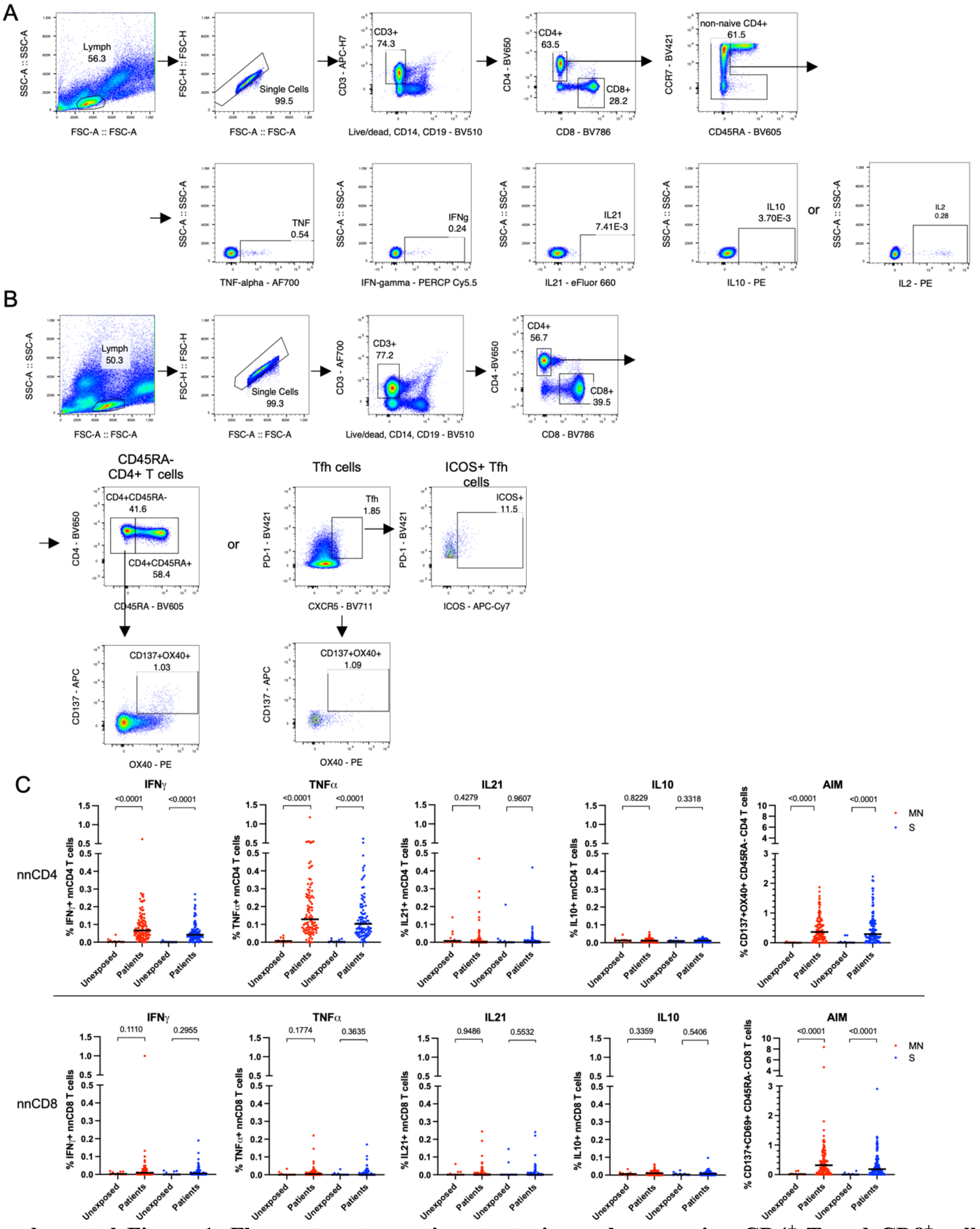

**Supplemental Figure 1. Flow cytometry gating strategies and comparing CD4⁺ T and CD8⁺ cell responses between unexposed controls and COVID-19 outpatients.**

Flow cytometry gating strategies for A) ICS or B) AIM assays are shown. C) PBMCs of unexposed controls (n=10) and COVID-19 outpatients at day 28 post-enrollment were stimulated with MN (ICS: n=102, AIM:

n=107) and S proteins (ICS: n=100, AIM=105) *in vitro*, stained and analyzed by flow cytometry. The absolute percentage of IFN$\gamma^+$, TNF$\alpha^+$, IL21$^{+,}$ IL10$^+$ non-naïve and AIM$^+$ CD45RA$^-$ CD4$^+$ T cells (nnCD4, top panel) CD4$^+$ T cells and CD8$^+$ T cells (nnCD8, bottom panel) post antigen stimulation (MN: red, S: blue) are illustrated (black line = median, Mann Whitney test).

### Supplemental Figure 2

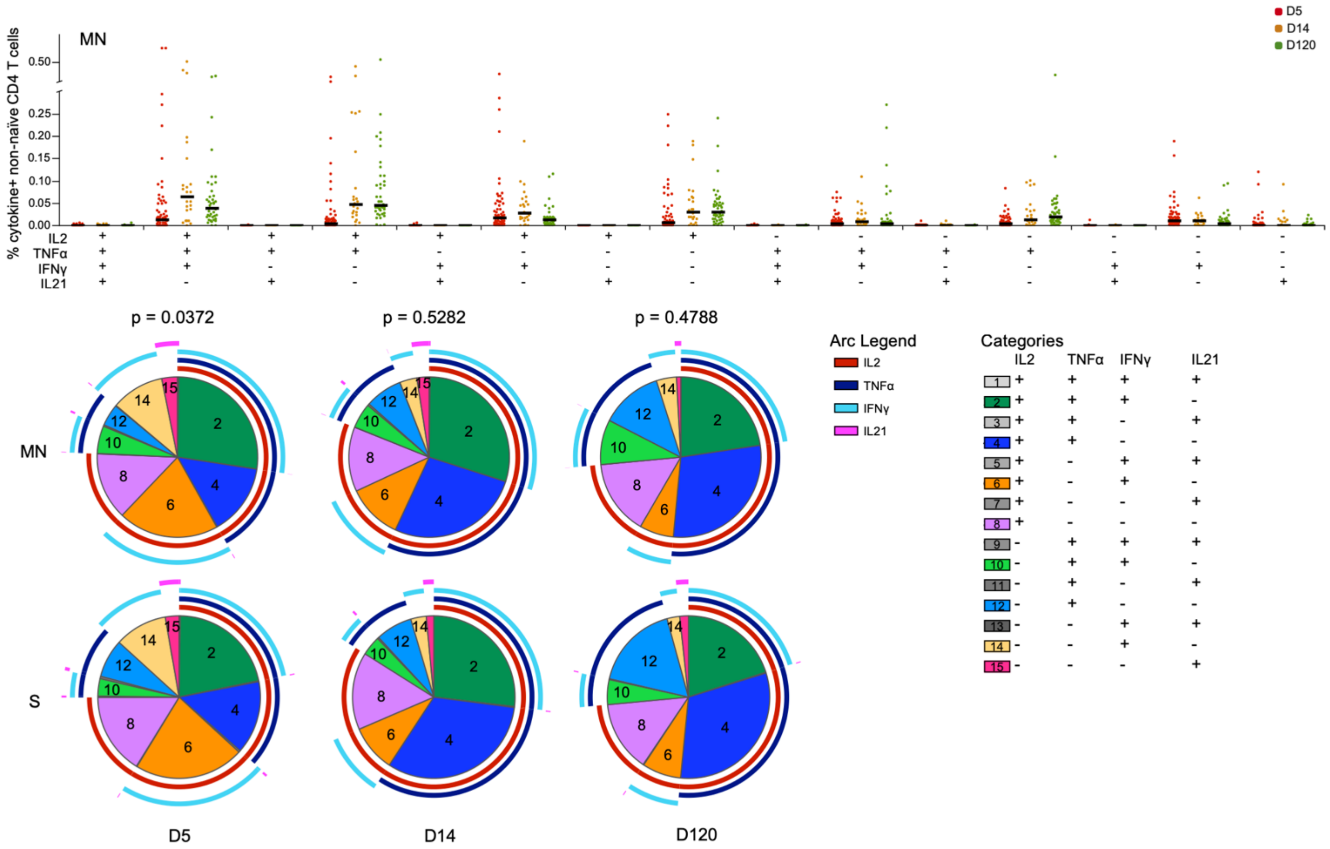

**MN**

| | | | |
|---|---|---|---|
| **D5** | X | | |
| **D14** | <0.0001 | X | |
| **D120** | <0.0001 | 0.0279 | X |
| **days** | **D5** | **D14** | **D120** |

**S**

| | | | |
|---|---|---|---|
| **D5** | X | | |
| **D14** | 0.0018 | X | |
| **D120** | <0.0001 | 0.0725 | X |
| **days** | **D5** | **D14** | **D120** |

**Supplemental Figure 2. The majority of IFNγ-, TNFα- or IFNγ- and TNFα-producing non-naïve CD4 T cells co-produce IL2.**

A) PBMCs of COVID-19 outpatients were stimulated with MN or S proteins in vitro, stained and analyzed by flow cytometry. Depicted are the absolute percentage (scatter plot, black line = median) and the relative proportion (pie charts, mean) of each individual combination of IL2, TNFα, IFNγ and IL21-producing non-naïve $CD4^{+}$ T cells (Boolean gating) stratified by days post-enrollment (D5: n=24, D14: n=24, D120: n=24). The P values indicated on top of the pie charts are calculated using the partial permutation test which tests the association between MN and S protein stimulation. The P values in the tables indicates the significance of the associations between the different days since symptom onset calculated using the partial permutation test (left = MN-stimulated T cells, right = S-stimulated T cells). Values shown are background ('media only' condition) subtracted.

#### Supplemental Figure 3

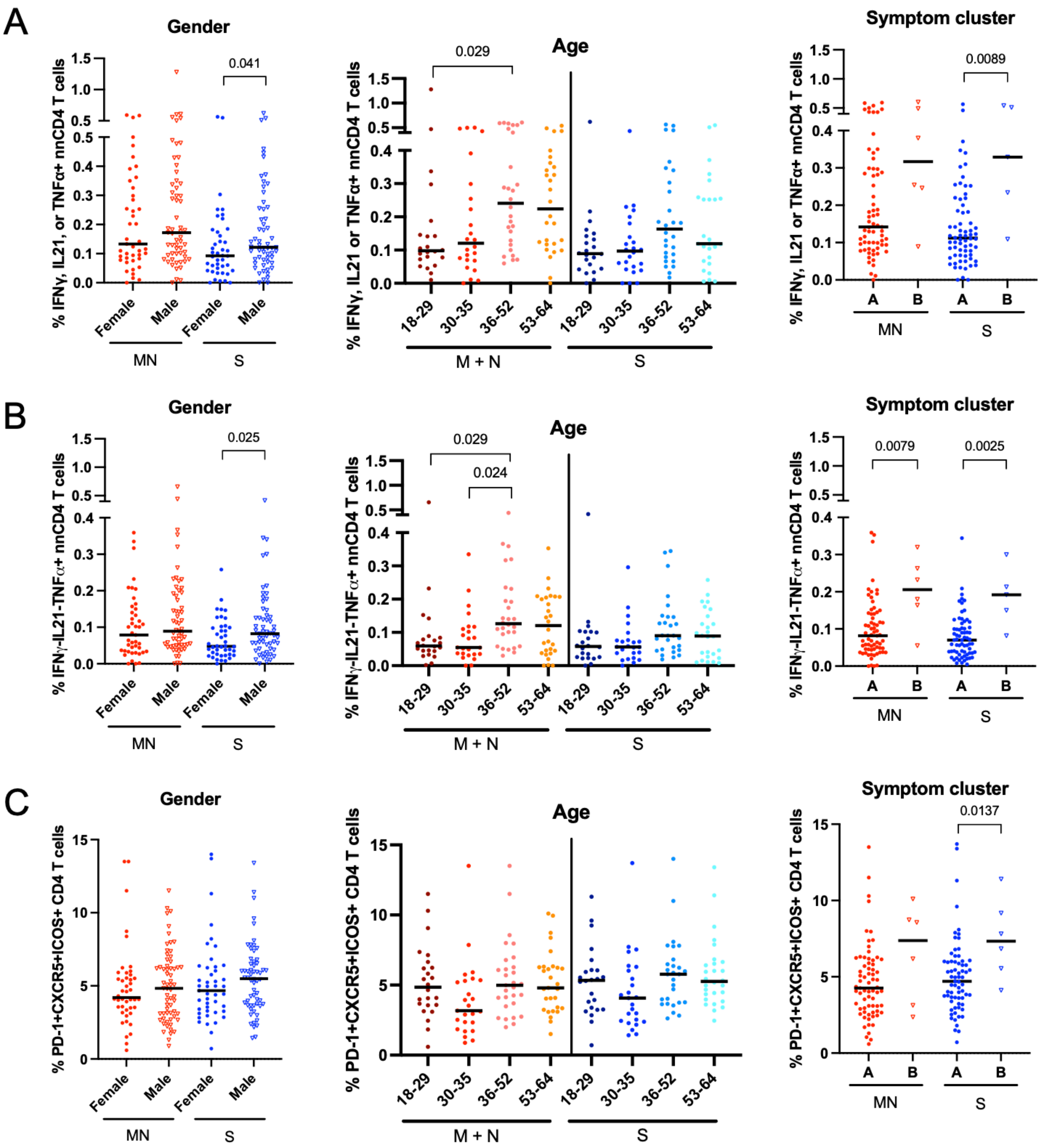

**Supplemental Figure 3 Day 28 IFN$\gamma^-$IL21$^-$TNF$\alpha^+$ non-naïve CD4$^+$ T cells are associated with gender, age and symptom cluster.**

Shown are scatter plots of absolute percentages of MN protein- (red) or S protein-stimulated (blue) A) IFNγ, IL21 or TNFα-producing non-naïve CD4$^+$ T cells, B) IFN$\gamma^-$IL21$^-$TNF$\alpha^+$ non-naïve CD4 T cells and C) ICOS$^+$ Tfh cells collected at day 28 post-enrollment (black line = median). The plots are stratified by gender (left), age (middle) and symptom cluster (right). Age categories were defined by participant age quartiles. Cluster A are characterized by higher prevalence of runny nose and sore throat. Cluster B are characterized by greater symptom severity and/or later peak severity (chest pain/pressure, fatigue, and myalgias) as described by Jacobson et al. P values shown in the T cells data versus gender and symptom cluster are calculated using the Mann Whitney test. Kruskal-Wallis rank sum test with Dunn's multiple comparisons test was used to calculate the P values in the T cell data versus age plots.

#### Supplemental Figure 4

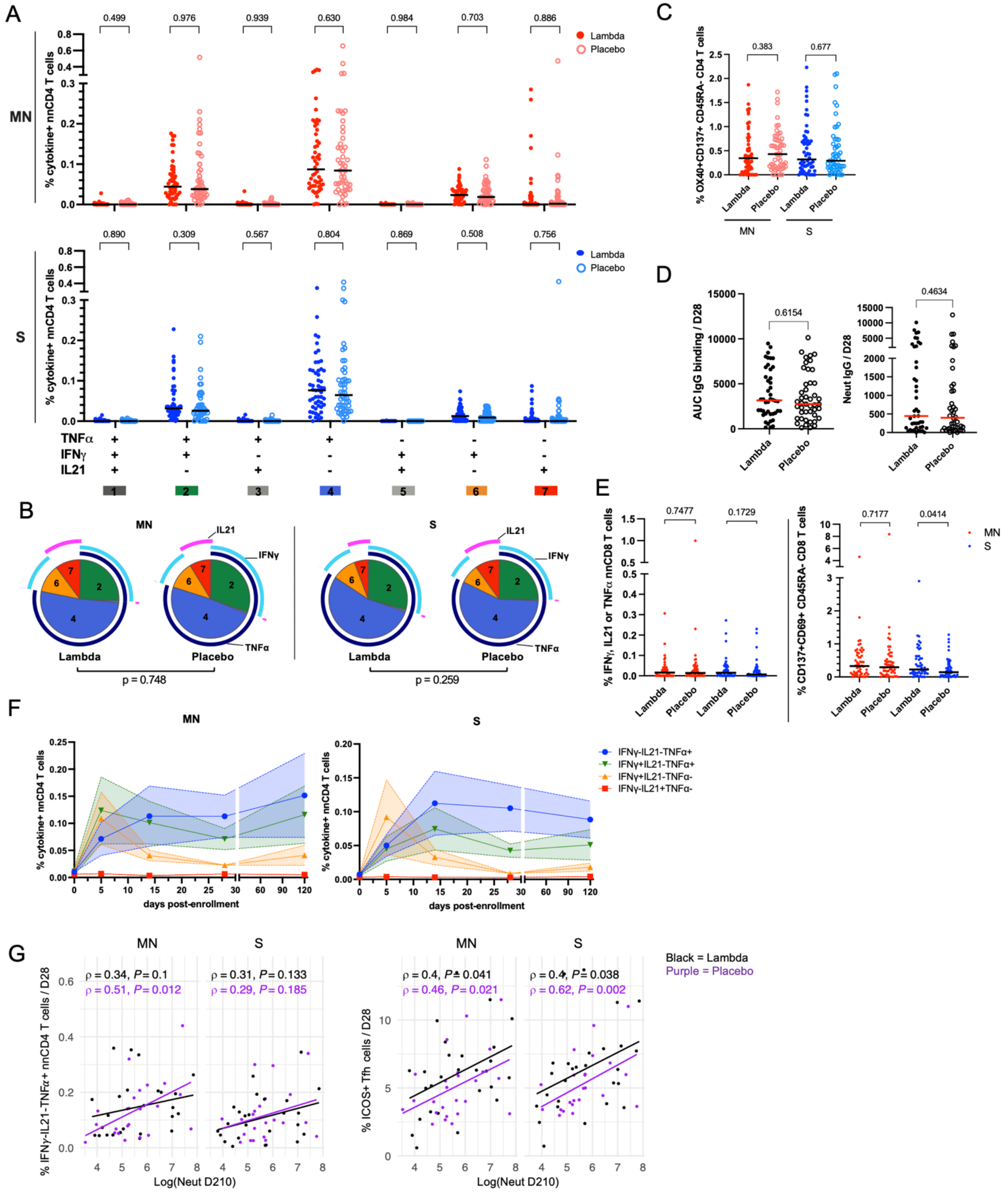

**Supplemental Figure 4. SARS-CoV-2-specific CD4$^{+}$ T cell subsets do not differ between participants randomized to Lambda or placebo.** PBMCs of COVID-19 outpatients on D28 post-enrollment were stimulated with spike (S) or combination of membrane (M) and nucleocapsid (N) proteins in vitro, stained and analyzed by flow cytometry. A) The absolute percentage (scatter plot, black line = median) of each individual combination of TNFα, IFNγ and IL21-producing non-naïve $CD4^+$ T cells stratified by treatment arms (closed circle: Lambda n=52, open circle: Placebo, n=50). Top panel depicts MN protein-stimulated $CD4^+$ T cells (red) and bottom panel S protein-stimulated $CD4^+$ T cells (blue). B) The pie charts represent the relative proportion (mean) of each individual combination of cytokine-producing non-naïve $CD4^+$ T cells are shown stratified by antigenic stimuli (MN n=102, S n=100). The P value shown under the pie charts is calculated using a partial permutation test. C) Shown are absolute percentages of $OX40^+CD137^+$ $CD45RA^-$ $CD4^+$ T cells stratified by treatment arms (Lambda n=54, Placebo n=53) and grouped by antigenic stimuli with the black line indicating the median. D) SARS-CoV-2 full length S binding IgG (AUC) (Lambda n=44, Placebo n=45) and neutralizing (Neut) antibody IgG titers (Lambda n = 44, Placebo n = 42) on D28 stratified in treatment arms are shown (red line = median). E) Stratified by treatment arm, in the top panel the absolute percentage of all $cytokine^+$ ($TNFa^+$ or $IFNg^+$ or $IL21^+$) non-naïve (left) or $AIM^+$ $CD45RA^-$ (right) $CD8^+$ T cells are depicted (placebo: n=50, Lambda: n=52, Black line = median). A, C-E) P values shown are calculated using the Mann Whitney test. F) PBMCs from 12 COVID-19 outpatients randomized to placebo arm sampled at day 5 (D5), day 14 (D14), day 28 (D28) and month 4 (D120) (n= 5 also sampled on day of enrollment (D0), D14: n=11 and D120: n=11) were stimulated with MN (left) or S (right) proteins in vitro, stained and analyzed by flow cytometry. The mean and SEM of the absolute percentage of background-subtracted single positive TNFα-, (blue), IFNγ- (yellow), IL21- (red) or TNFα and IFNγ-producing non-naïve $CD4^+$ T cells (green) of samples derived from the placebo arm only at sequential time points post-enrollment are shown. G) Scatter plots comparing antigen-stimulated $IFN\gamma^-IL21^-TNF\alpha^+$ non-naïve $CD4^+$ T cells (MN: n=48, S: n=47) or $ICOS^+$ Tfh cells (right, MN: n=52 , S: n=50) collected at D28 with neutralizing antibody titers collected at day 210 (D210) post-enrollment are shown, which are stratified in treatment arms (Lambda = black, Placebo = purple). The neutralizing antibody titers are presented in natural logarithm and added +1 to allow for inclusion of participants with no neutralizing activity. The rho (ρ) and P values were calculated using spearman's correlation (Benjamini-Hochberg corrected). The lines represent the fitted linear relationship between the indicated data. All values shown are background ('media only' condition) subtracted. AUC = area under the curve.

#### Supplemental Figure 5

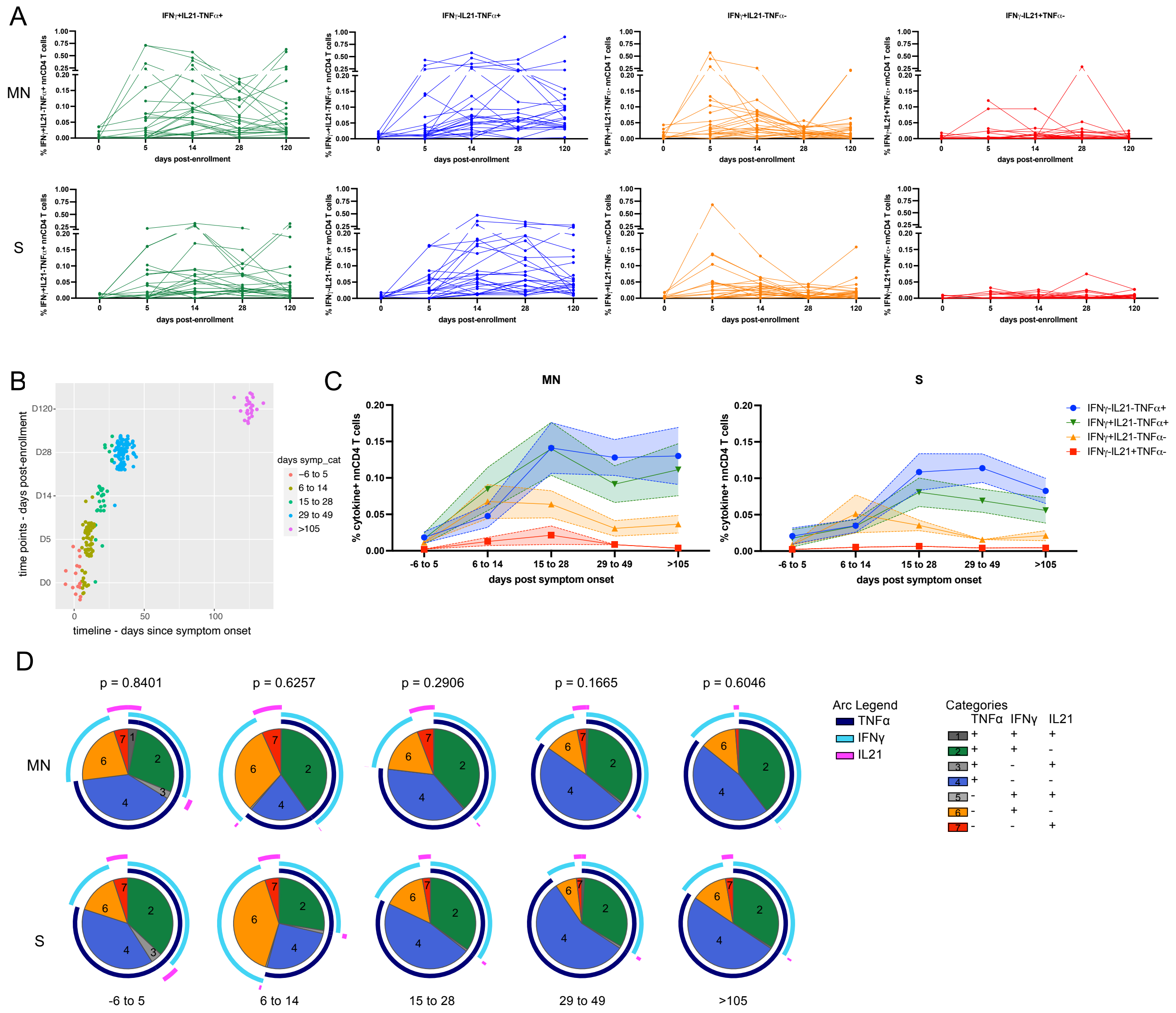

**MN**

| | | | | | |
|---|---|---|---|---|---|
| -6 - 5 | X | | | | |
| 6 - 14 | 0.0123 | X | | | |
| 15 - 28 | 0.0056 | 0.0089 | X | | |
| 29 - 49 | 0.0077 | <0.0001 | 0.0120 | X | |
| >105 | 0.0003 | <0.0001 | 0.0011 | 0.4763 | X |
| days | -6 - 5 | 6 - 14 | 15 - 28 | 29 - 49 | >105 |

**S**

| | | | | | |
|---|---|---|---|---|---|
| -6 - 5 | X | | | | |
| 6 - 14 | 0.6210 | X | | | |
| 15 - 28 | 0.1942 | 0.0049 | X | | |
| 29 - 49 | 0.0002 | <0.0001 | 0.0030 | X | |
| >105 | 0.0179 | 0.0002 | 0.0749 | 0.4002 | X |
| days | -6 - 5 | 6 - 14 | 15 - 28 | 29 - 49 | >105 |

**Supplemental Figure 5. Longitudinal analysis of quality and magnitude of SARS-CoV-2-specific $CD4^+$ T cell responses over time.**

A) Shown are absolute percentages of paired individual values of indicated non-naïve $CD4^+$ T cell populations which are connected over time post-enrollment. B) Patients were re-grouped based on days since symptom onset (-6 to 5 (n=16), 6 to 14 (n=67), 15 to 28 (n=30), 29 to 49 (n=94) and >105 days (n=23)). Symptom data was missing for 6 patients, resulting in 10 missing data points in the days since onset group. The graph depicts the distribution of days since symptom onset on a timeline (x-axis) versus the grouped days post-enrollment (y-axis). The new categories defined are shown in different colors. The categories are selected with the kinetics of antigen-specific T cell development in mind. The numbers were chosen based on number of weeks (-6 to 5 = <1 week, 6 to 14 = < or =2 weeks, 15 to 28 = 2 – 4 weeks, 29 to 49 = 4 to 7 weeks, >105 = >15 weeks). C) The mean and SEM of the absolute percentage of background-subtracted $IFN\gamma^{-}IL21^{-}TNF\alpha^{+}$ (blue), $IFN\gamma^{+}IL21^{-}TNF\alpha^{-}$ (yellow), $IFN\gamma^{-}IL21^{+}TNF\alpha^{-}$ (red) or $IFN\gamma^{+}IL21^{-}TNF\alpha^{+}$ non-naïve $CD4^{+}$ T cells (green) of sequential timepoints of days since symptom onset are shown. Non-naïve $CD4^{+}$ T cells stimulated with MN (lef) or S (right) proteins are illustrated. D) Depicted are the relative proportion (pie charts, mean) of each individual combination of TNFα, IFNγ and IL21-producing non-naïve $CD4^{+}$ T cells (Boolean gating) stratified by days since symptom onset are shown (-6 to 5: n=8, 6 to 14: n=26, 15 to 28: n=23, 29 to 49: n=23, and >105: n=23). The P values indicated on top of the pie charts are calculated using the partial permutation test which tests the association between MN and S protein stimulation. The P values in the tables indicates the significance of the associations between the different days since symptom onset calculated using the partial permutation test (left = MN protein-stimulated T cells, right = S protein-stimulated T cells). Values shown are background ('media only' condition) subtracted.

#### Supplemental Figure 6

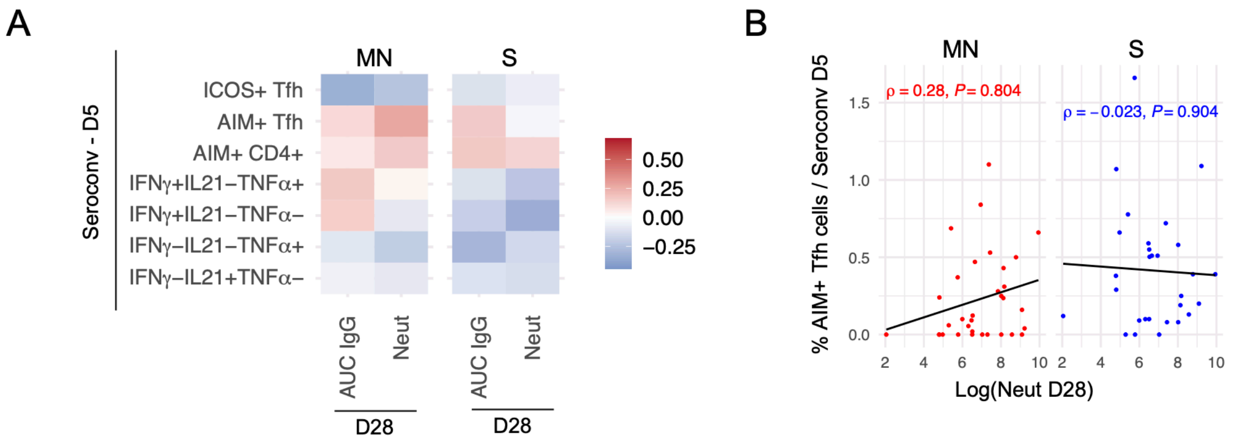

**Supplemental Figure 6. Early T cell responses of participants who seroconverted do not correlate with late SARS-CoV-2-specific antibody titers.** A) In the heatmap spearman's correlations (Benjamini-Hochberg corrected) are shown between the indicated MN protein- (left) or S protein-stimulated (right) T cell data collected at day 5 (D5) from participants who seroconverted and S protein IgG binding (AUC IgG) or neutralizing antibody (Neut) titers collected at day 28 (D28) post-enrollment. Participants who seroconverted (n=39) were defined by lacking S protein binding IgG titers at the start of enrollment and having >0.25 $OD_{450}$ collected by ELISA at later timepoints. B) Scatter plots comparing MN protein- (left/red) and S protein-stimulated (right/blue) $AIM^{+}$ Tfh cells collected at D5 from participants who seroconverted with neutralizing titers at D28 (Neut D28) post-enrollment are depicted (MN: n=33, S: n=29). The neutralizing antibody titers are presented in natural logarithm and added +1 to allow for inclusion of participants with no neutralizing activity. The rho (ρ) and P values were calculated using spearman's correlation (Benjamini-Hochberg corrected). The lines represent the fitted linear relationship between the indicated data. AUC = area under the curve.

#### Supplemental Figure 7

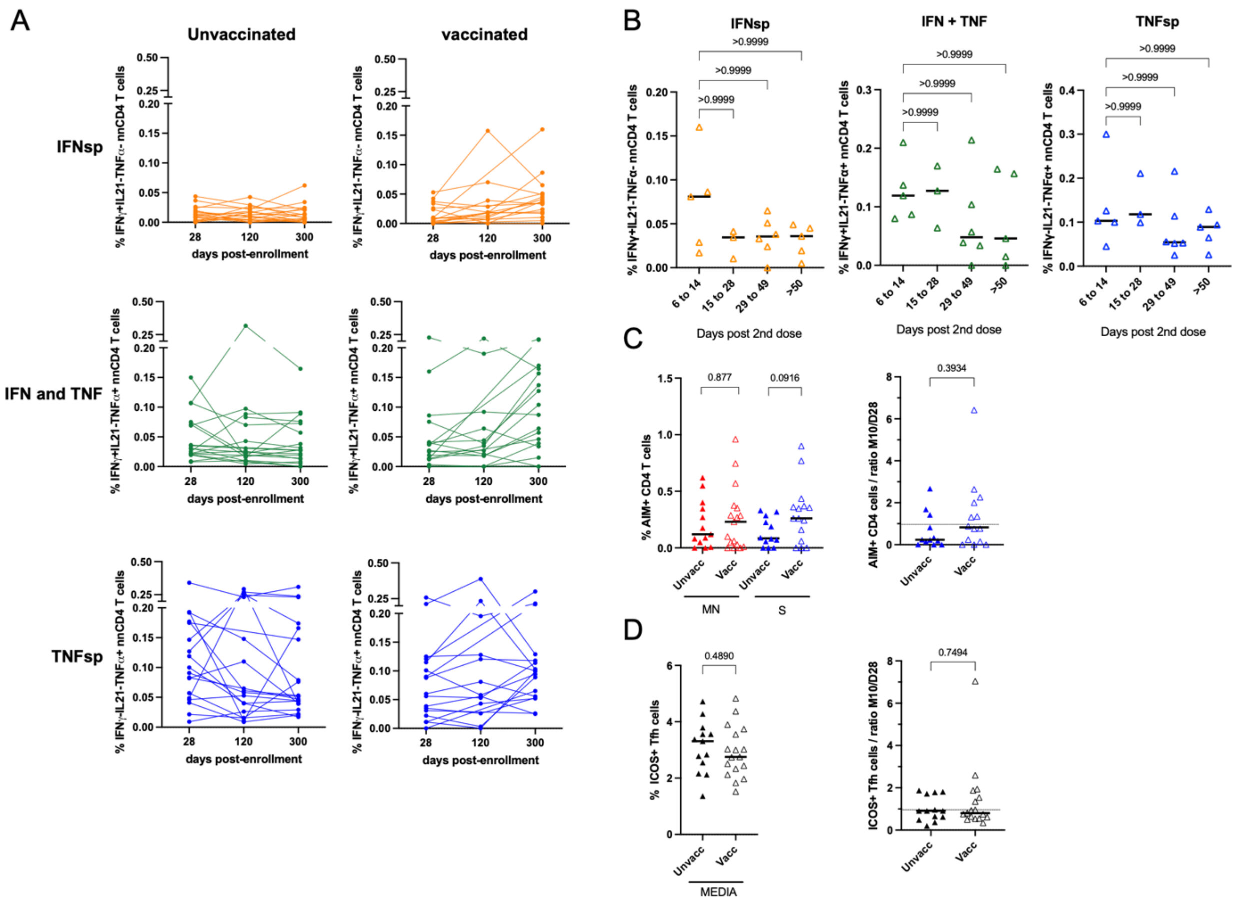

**Supplemental Figure 7. Magnitude of IFNγ$^+$IL21$^-$TNFα$^-$ and IFNγ$^+$IL21$^-$TNFα$^+$ non-naïve CD4$^+$ T cells following mRNA vaccination increases compared to D28 and M4 time points.** PBMCs of COVID-19 outpatients who received full doses of the mRNA COVID-19 vaccine and who did not receive the vaccine prior to the M7 or M10 follow-up visit were stimulated with MN or S proteins in vitro, stained and analyzed by flow cytometry. A) The absolute percentage of paired IFNγ$^+$IL21$^-$TNFα$^-$ (IFNsp, yellow), IFNγ$^+$IL21$^-$TNFα$^+$ (IFNγ and TNFα, green), and IFNγ$^-$IL21$^-$TNFα$^+$ non-naïve CD4$^+$ T cells (TNFsp, blue) after S protein stimulation of D28, D120 (M7) and D300 (M10) post-enrollment are shown (n=19). B) The absolute percentage of indicated cytokine-producing non-naïve CD4$^+$ T cells stratified by days since second dose at the time of sampling are depicted. The days categories are selected with the kinetics of antigen-specific T cell development in mind. The numbers were chosen based on number of weeks (6 to 14 = < or = 2 weeks, 15 to 28 = 2 – 4 weeks, 29 to 49 = 4 to 7 weeks, >50 = > 8 weeks). P values are calculated by using the Kruskal-Wallis rank sum test with Dunn's multiple comparisons test. C) Stratified by vaccination status, the absolute percentage (left) and the ratio between M10 and D28 of AIM$^+$ (CD137$^+$/OX40$^+$) CD45RA$^-$ CD4$^+$ T cells (right) stimulated with the MN (red) or S protein (blue) are shown. D) Stratified by vaccination status, the absolute percentage (left) and the ratio between M10 and D28 of unstimulated ('media only') ICOS$^+$ Tfh cells (PD-1$^+$CXCR5$^+$) (right) are shown. C, D) The P values shown are calculated using the Mann Whitney test.
